# Human Genetic Analysis Reveals Circulating Alpha-1 Antitrypsin Level as a Protective Factor in Sepsis

**DOI:** 10.64898/2026.03.25.26349312

**Authors:** Dandan Tan, Ping Zhang, Thomas M. Zheng, Kevin Y.H. Liang, Chen-Yang Su, Yiheng Chen, Tianyuan Lu, J Brent Richards, Amanda Y. Chong, Patrick R. Lawler, Fergus Hamilton, Alexander J. Mentzer, Julian C. Knight, Guillaume Butler-Laporte

**Author notes:** Corresponding author: Guillaume Butler-Laporte.

## Abstract

Sepsis is a dysregulated host response to infection and a leading cause of global mortality, yet effective targeted therapies remain lacking. Here, we applied a proteogenomic framework integrating large-scale human genetics with circulating proteomics to identify therapeutic targets. In a meta-analysis of genome-wide association studies of 60,314 sepsis cases and 1,464,733 controls, we identified four genome-wide significant loci, including a missense variant in *SERPINA1*, encoding alpha-1 antitrypsin (AAT), that was also associated with 30-day sepsis mortality in the UK Biobank.

Mendelian randomization (MR) and colocalization analyses supported a causal and protective effect of higher genetically predicted circulating AAT levels on sepsis risk. The protective association was highly specific to acute infectious phenotypes, including pneumonia, and was not observed for non-infectious traits. In two independent cohorts (UK Genomic Advances in Sepsis and the Biobanque Québécois sur la COVID-19), circulating AAT increased markedly during acute illness but was significantly attenuated among missense variant carriers in a dose-dependent manner, consistent with impaired protease-antiprotease balance.

MR of the AAT-regulated proteome recapitulated findings from prior sepsis trials, both negative and positive, providing orthogonal genetic support for therapeutic modulation of this pathway. Together, these findings provide the first human genetic evidence for AAT’s causal role in sepsis, positioning *SERPINA1* as a high-priority candidate for drug repurposing and targeted therapeutic interventions.

## Introduction

Sepsis is a critical medical condition caused by dysregulated immune response to infection, accounting for an estimated 20% of global mortality^1,2^. In the United States alone, sepsis contributes to over one-third of in-hospital deaths, with healthcare costs exceeding $38 billion in 2017^3^. Because the condition progresses rapidly, prompt and comprehensive clinical management is vital^4^; however, current treatment strategies remain limited to supportive care and antimicrobial therapy^5^. Despite decades of investigation, with the notable exception of increasing use of corticosteroids^6,7^, no new pharmacological treatments for sepsis have been approved in recent years, and many high-profile clinical trials have failed^8–11^.

This lack of progress reflects several fundamental challenges. First, sepsis is an “umbrella” term for a clinical syndrome arising from diverse infectious etiologies^12,13^. Second, its presentation can be highly heterogeneous, influenced by complex systemic inflammation and confounded by age, socioeconomic status, and comorbidities^13–15^. Third, the rapid clinical course complicates observational studies, in which reverse causation, whether biomarkers are drivers or consequences of disease, is difficult to resolve. While the initial clinical management of sepsis is often similar regardless of the causative pathogen, the discovery of effective therapies requires a more precise understanding of the shared biological drivers that transcend clinical heterogeneity.

To address this gap, we used a proteogenomic approach to identify and validate molecular drivers of sepsis. Because germline genetic variants are randomly assigned at conception and remain fixed throughout life, they provide a natural experiment to assess causality with minimal confounding and reduced risk of reverse causation, an approach known as Mendelian randomization (MR)^16^. By integrating large-scale genome-wide association studies (GWAS) with proteomic data, we can mitigate the pitfalls of traditional observational studies and highlight targets with high translational potential^17,18^.

In this study, we conducted a genome-wide meta-analysis of 60,314 cases and 1,464,733 controls, identifying four significant loci (P < 5 × 10^-8^) with consistent effect sizes across cohorts (Fig. 1). We prioritized a well-known missense variant in *SERPINA1*. This gene encodes alpha-1 antitrypsin (AAT), a potent serine protease inhibitor that regulates the inflammatory response by neutralizing neutrophil elastase (ELANE) and other proteases^19,20^. We then performed multiple downstream analyses to support its potential as a therapeutic target, including mortality analyses, MR analyses on multiple disease categories, observational validation of circulating AAT in two independent cohorts (UK Genomic Advances in Sepsis [UK GAinS]^21^ and The Biobanque Québécois sur la COVID-19 [BQC19]^22^), and comparison between MR of AAT and other proteins with previously published randomized trials.

**Figure 1.**
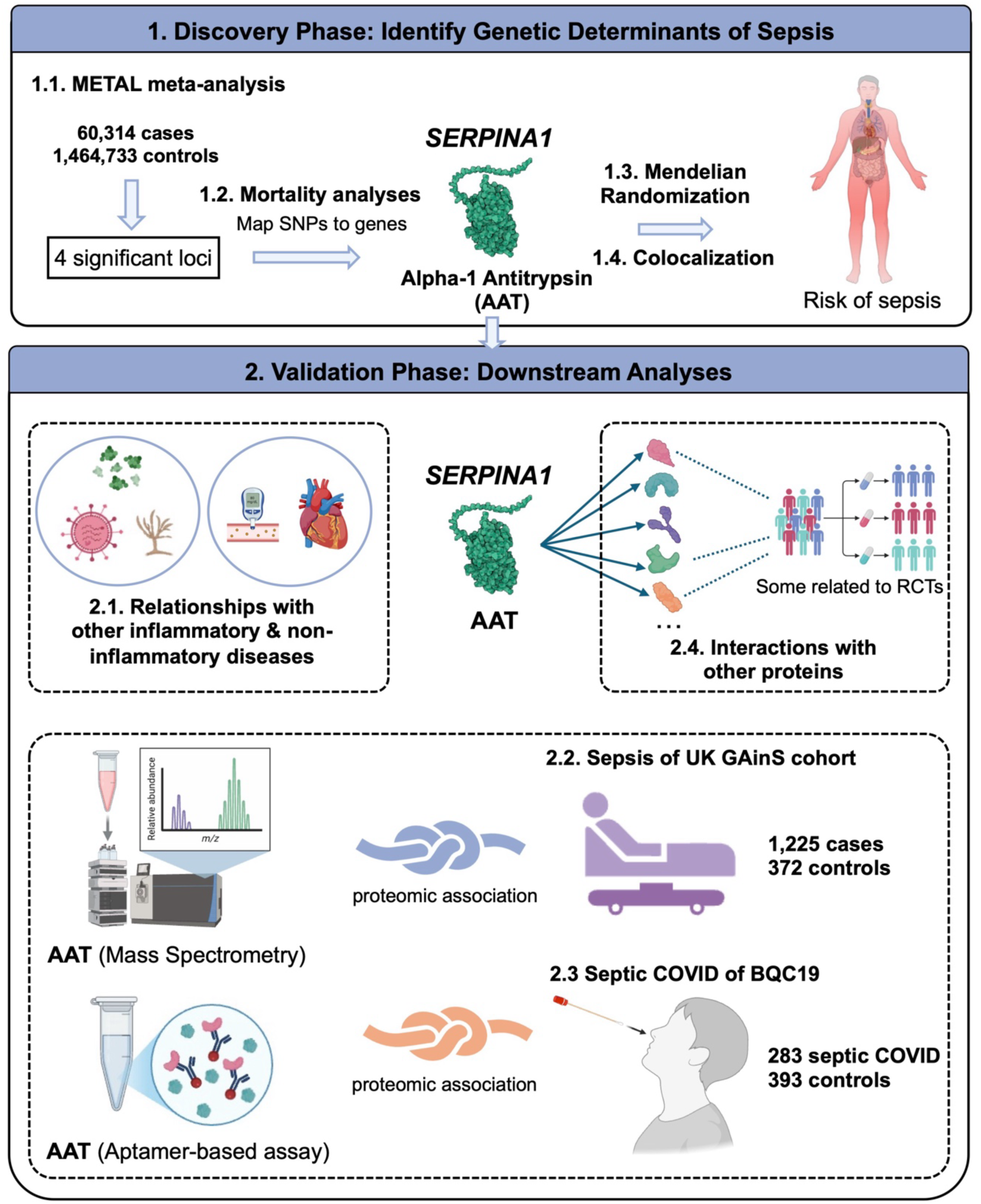
Proteogenomic workflow for the identification and validation of *SERPINA1* and its encoded protein alpha-1 antitrypsin as a therapeutic target in sepsis. Overview of the multi-stage study design integrating large-scale genomic data with clinical and proteomic validation. The discovery phase consisted of a genome-wide association meta-analysis of sepsis across four cohorts (UK Biobank, FinnGen, Million Veteran Program, and All of Us), totaling 60,314 cases and 1,464,733 controls, which identified four genome-wide significant loci (P < 5 × 10^-8^). Following mortality analyses to assess clinical relevance, the *SERPINA1* locus on chromosome 14 was prioritized based on a lead missense variant. Causal inference was performed using two-sample Mendelian randomization (MR) and colocalization, utilizing alpha-1 antitrypsin (AAT) protein quantitative trait loci (pQTLs) as the exposure to determine the effect of circulating AAT levels on sepsis risk. In the validation phase, the functional profile of AAT was further explored via MR across a broad phenome, including infection-related conditions (e.g., pneumonia, bronchiectasis, skin infections, and urinary tract infections) and metabolic traits (e.g., type 2 diabetes, hypertension), also some known AAT-related traits (e.g., cirrhosis, chronic obstructive pulmonary disease(COPD)). The workflow concludes with observational validation of AAT protein dynamics in two independent sepsis cohorts (UK GAinS and BQC19) and an exploratory MR analysis between AAT and other protein, including those targeted in previous randomized controlled trials, to evaluate therapeutic potential.

Overall, evidence triangulation from genetic variation, MR, colocalization, and proteomics supports a central role for *SERPINA1* and its corresponding protein, AAT, in sepsis pathogenesis and highlights its potential for therapeutic development.

## Results

### Meta-analysis identifies four genome-wide significant loci for sepsis

To identify genetic drivers of sepsis, we performed a fixed-effects meta-analysis using METAL^23^ across four large-scale cohorts: UK Biobank (UKB)^24^, Million Veteran Program (MVP)^25^, FinnGen^26^, and All of Us (AoU)^27^. This combined dataset comprised 60,314 sepsis cases and 1,464,733 controls of European ancestry (Fig. 2a, Supplementary Table 1). Cohort-specific sepsis definitions are described in the Methods. Four independent loci reached genome-wide significance on genome build 38, each demonstrating consistent effect directions across the contributing cohorts (Fig. 2b, Supplementary Table 2).

**Figure 2.**
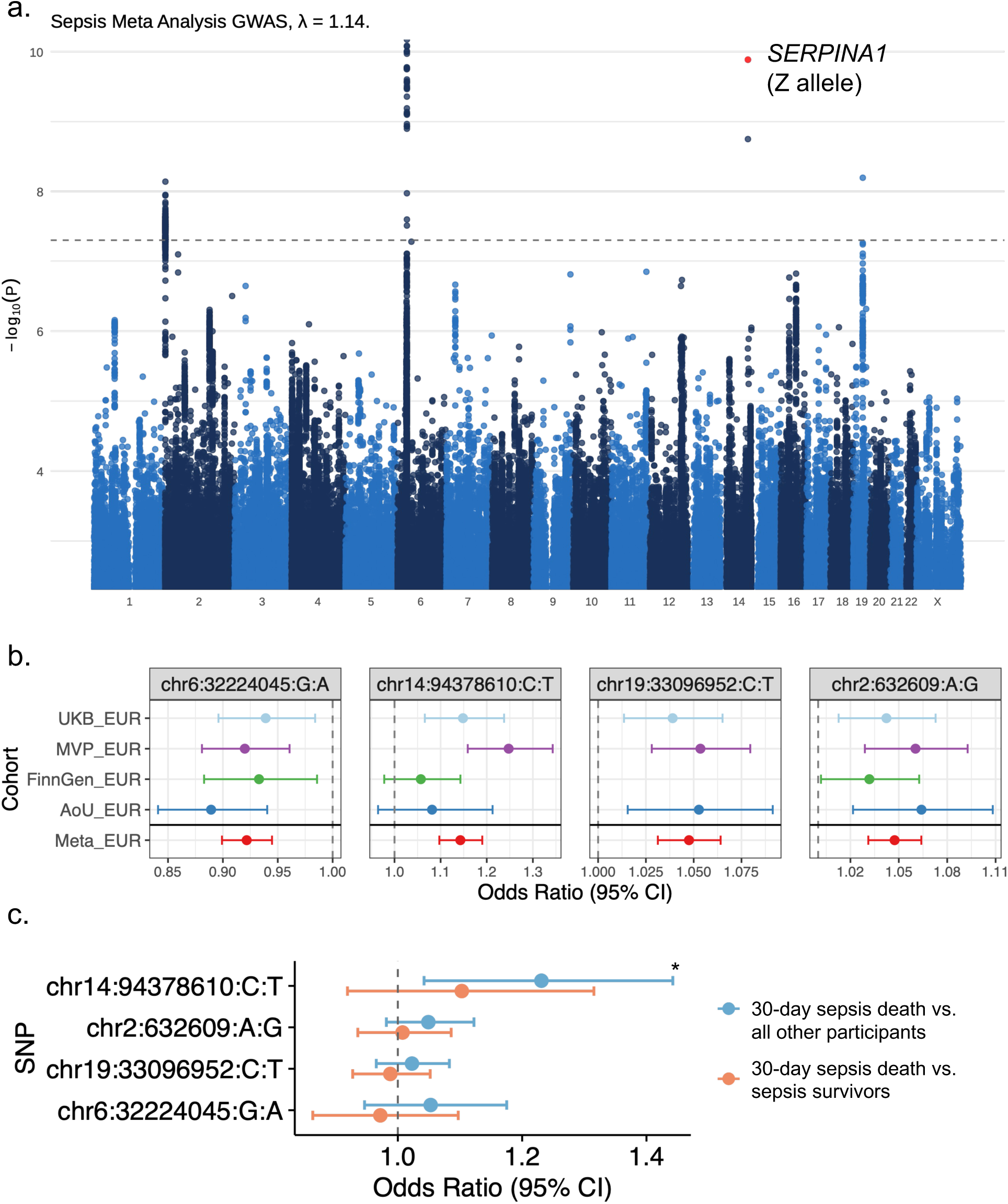
Genome-wide discovery and clinical relevance of the *SERPINA1* locus. a) Manhattan plot illustrating the meta-analysis of genetic associations with sepsis (n = 1,525,047). The x-axis denotes chromosomal position, and the y-axis represents the negative of the log_10_(P) value (the higher the dot, the lower the p-value). The horizontal dashed line indicates the threshold for genome-wide significance (P = 5 × 10^-8^). The genomic inflation factor (λ) was 1.14. Red dot on the plot highlights the *SERPINA1* missense variant (the Z allele). The atypical, narrow peak architecture at this locus reflects the limited linkage disequilibrium (LD) of the Z allele, which is in high LD (R^2^ = 0.92) with only one other variant (chr14:94371805:G:T). b) Forest plot demonstrating the consistency of effect directions for the four genome-wide significant variants across the four discovery cohorts (UKB, MVP, FinnGen, and AoU). The chromosome 19 variant is excluded in FinnGen after genotype quality control. c) Mortality analysis in the UK Biobank assessing the association between sepsis risk alleles dosage and 30-day sepsis mortality. Points represent odds ratios (OR) and error bars denote 95% confidence intervals (CI). The blue line represents the association using a 30-day sepsis death compared with all remaining UKB individuals, reflecting overall risk of septic death. The red line represents a subgroup analysis restricted to sepsis cases, where we compared 30-day sepsis death versus sepsis survivors beyond 30 days, specifically evaluating the effect of the variant on disease severity. * means statistically significant (P < 0.05). UKB_EUR, UK Biobank European ancestry; MVP_EUR, Million Veteran Program European ancestry; FinnGen_EUR, FinnGen European ancestry; AoU_EUR, All of Us Research Program European ancestry; Meta_EUR, meta-analysis of the four European-ancestry discovery cohorts.

The most statistically significant signal mapped to the human leukocyte antigen (HLA) region (chr6:32224045:G:A; OR = 0.92, 95% CI = 0.90-0.94, P = 1.0 × 10^-10^), aligned with the established immune-mediated architecture of sepsis susceptibility. A second locus was identified on chromosome 14 within the *SERPINA1* gene (chr14:94378610:C:T; OR = 1.14, 95% CI = 1.10-1.19, P = 1.3 × 10^-10^); given its high biological plausibility, large effect size, and clinical relevance, this locus became the primary focus of our downstream investigations. On chromosome 19, a third signal (chr19:33096952:C:T; OR = 1.05, 95% CI = 1.03-1.06, P = 6.4 × 10^-9^) was identified near to *GPATCH1* (Supplementary Fig. 1), which encodes a spliceosome-associated protein that interacts with the innate immune factor DHX35^28,29^. Finally, a fourth significant signal was identified on chromosome 2, a third (chr2:632609:A:G; OR = 1.05, 95% CI = 1.03-1.06, P = 7.2 × 10^-9^) was located about 30 kilobases from *TMEM18*, a gene previously implicated in obesity risk^30^ (Supplementary Fig. 2).

### Prioritization of a *SERPINA1* missense variant

For the remainder of this paper, we focused on the chromosome 14 locus (chr14:94378610:C:T), which harbored a missense variant in *SERPINA1*. This gene encodes AAT, a hepatocyte-produced serine protease inhibitor that protects tissues, most notably the lungs, from protease-mediated inflammatory damage^31^. Pathogenic mutations in *SERPINA1* are the monogenic cause of alpha-1 antitrypsin deficiency (AATD)^32^.

Historically, *SERPINA1* alleles have been classified by their electrophoretic migration patterns: the wild-type allele is designated “M”, while variants migrating proximal or distal to the wild-type allele are labeled A-L and N-Z, respectively^33^. The lead variant identified in our meta-analysis corresponds to the Z allele (Glu342Lys), which underlies the most clinically severe form of AATD. In addition, this pathogenic variant is highly stratified by population, maintaining an allele frequency of approximately 2% in European ancestries while being nearly undetectable in non-European populations. Individuals homozygous for this allele (PI*ZZ) possess only 10-20% of normal circulating AAT levels and are at high risk of early-onset emphysema and cirrhosis^31,34^. Heterozygotes (PI*MZ) exhibit mild-to-moderate protein reductions, typically maintaining around 60% of normal AAT levels^33^. The Z mutation creates a dual pathological challenge: the lack of protective AAT in the circulation allows protease like ELANE to degrade lung tissue, while the accumulation of misfolded AAT within hepatocytes triggers liver inflammation and damage^35,36^.

In the sepsis meta-analysis, we observed a second independent missense variant of *SERPINA1* (chr14:94380925:T:A; OR = 1.04, 95% CI = 1.01-1.08, P = 9.8 × 10^-3^), corresponding to the S allele (Glu264Val; allele frequency = 5% in Europeans). Compared to the Z allele, the S variant confers a milder reduction of AAT and correspondingly lower risk of lung or liver pathology, as the S mutation has a less pronounced impact on protein conformation and secretion^37^. The fact that both Z and S alleles are independently associated with sepsis risk, with the magnitude of the effect mirroring the severity of the protein deficiency, strongly identifies *SERPINA1* as the likely effector gene at this locus. Given that AAT is a primary inhibitor of ELANE via the formation of an irreversible complex^38,39^, we hypothesized that genetically or physiologically elevated AAT levels protect against sepsis by regulating systemic neutrophil protease activity.

### *SERPINA1* missense variant increases risk of sepsis mortality

To assess clinical impact of the identified loci, we examined the association between genotype dosage (0/1/2) of these risk alleles and 30-day sepsis mortality in the UKB (defined as death occurring between five days prior to and 30 days following diagnosis using UKB hospital records). Among the four genome-wide significant loci, the *SERPINA1* missense variant demonstrated the strongest association with mortality.

Compared to the general population, carriers of the *SERPINA1* Z allele had a significantly higher risk of 30-day sepsis mortality (3,131 cases; 457,750 controls) (OR = 1.23, 95% CI = 1.04-1.44, P = 0.01) (Fig. 2c, Supplementary Table 3). To ensure this finding was not just a reflection of increased sepsis susceptibility, we further compared sepsis patients who died within 30 days (n = 3,131) specifically against sepsis survivors (n = 14,334). Because the Z allele strongly increases sepsis incidence, evaluating mortality exclusively among sepsis patients introduces the risk of collider bias. This phenomenon typically attenuates genetic associations toward the null, potentially masking a true effect. Despite this expected downward bias, the direction of association in our cases-only subgroup remained consistently linked to increased mortality risk (OR = 1.10, 95% CI = 0.92-1.32). Although it did not reach formal statistical significance (P = 0.28), likely due to the reduced sample size of the sepsis-only cohort, the stability of effect size suggests that the Z allele contributes to mortality risk independently of its role in sepsis acquisition.

### Mendelian randomization and colocalization indicate alpha-1 antitrypsin (AAT) protection

To establish a causal link between circulating protein levels and sepsis risk, we performed two-sample MR. We utilized *cis*-pQTLs from two large-scale proteomic studies, deCODE^40^ and UK Biobank Pharma Proteomics Project (UKB-PPP)^41^, as genetic instruments for AAT levels. These analyses demonstrated that genetically predicted higher AAT concentrations significantly reduced sepsis risk (OR = 0.81, P = 1.4 × 10^-11^; UKB-PPP) (Fig. 3). Colocalization analyses supported these findings, indicating a shared causal variant between the genomic signals for protein expression and disease risk (PP.share = 99%) (Extended Data Fig. 1).

**Figure 3.**
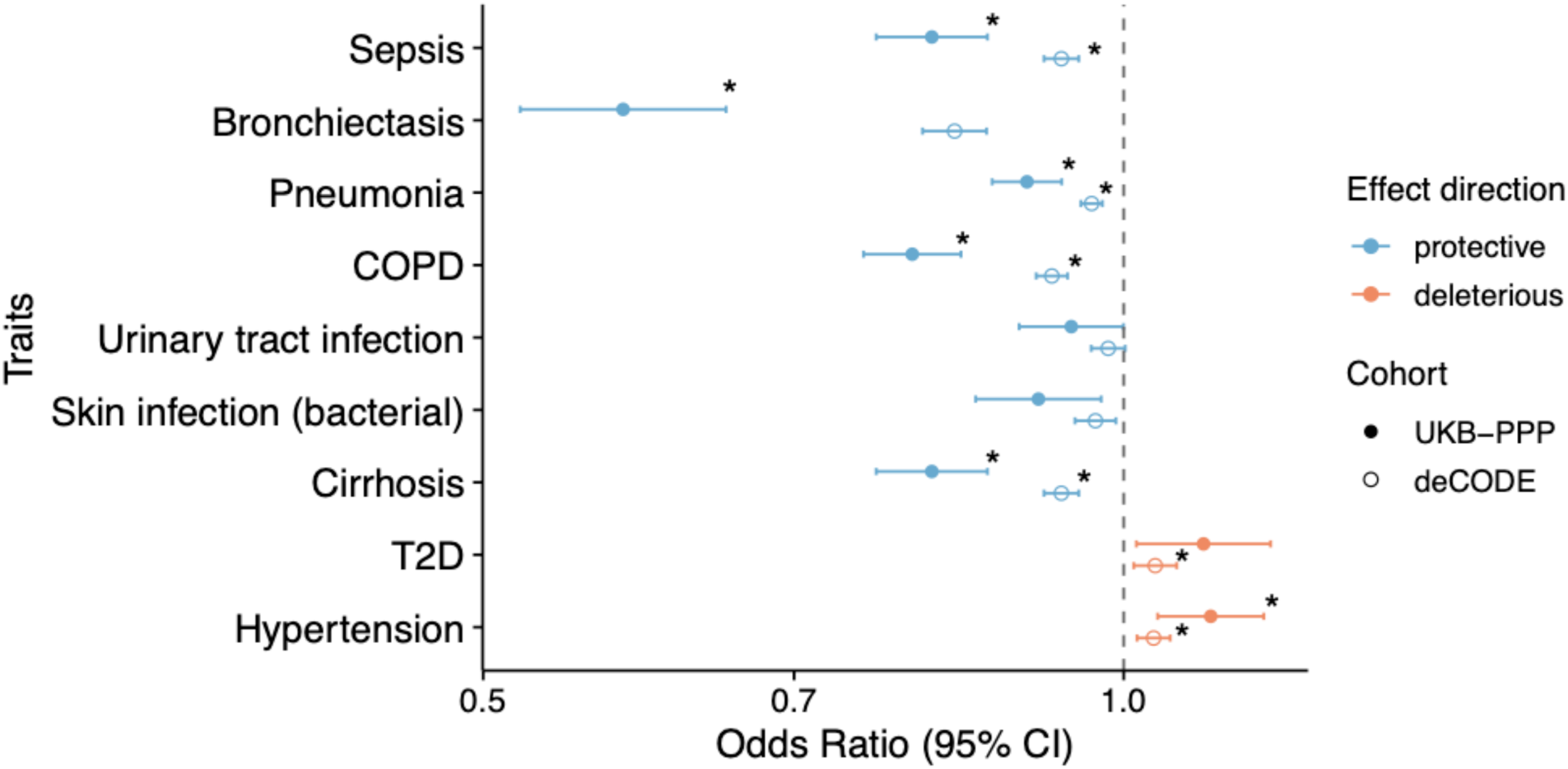
Causal evidence for AAT in sepsis and related phenotypes. Mendelian randomization (MR) estimates illustrating the causal effect of genetically predicted circulating AAT levels on sepsis risk and diverse clinical outcomes. Results are derived using independent pQTL instruments from both the UKB-PPP and deCODE datasets. Points represent odds ratios per standard deviation increase in protein levels, with error bars denoting 95% confidence intervals (CI) using fixed-effects inverse variance weighted method. Blue indicates a protective effect (OR < 1), while red indicates a deleterious association (OR > 1). Traits include respiratory infections (bronchiectasis, pneumonia), systemic infections (urinary tract and bacterial skin infections), lung and liver-related disease (COPD, cirrhosis), and chronic metabolic conditions (T2D, and hypertension). * represents statistically significant after Bonferroni adjustment (P < 0.005, which is 0.05 / 9).

Our results further revealed a quantitative genetic dose-response relationship. The Z allele, which is associated with the most significant reduction in circulating AAT levels (β =-0.50), conferred a higher risk of sepsis than the S allele, which is more common but results in a less protein reduction (β =-0.37) (Fig. 4a). This dosage-dependent relationship between protein levels and disease risk provides powerful evidence that AAT acts as a causal driver of sepsis susceptibility, rather than a mere biomarker.

**Figure 4.**
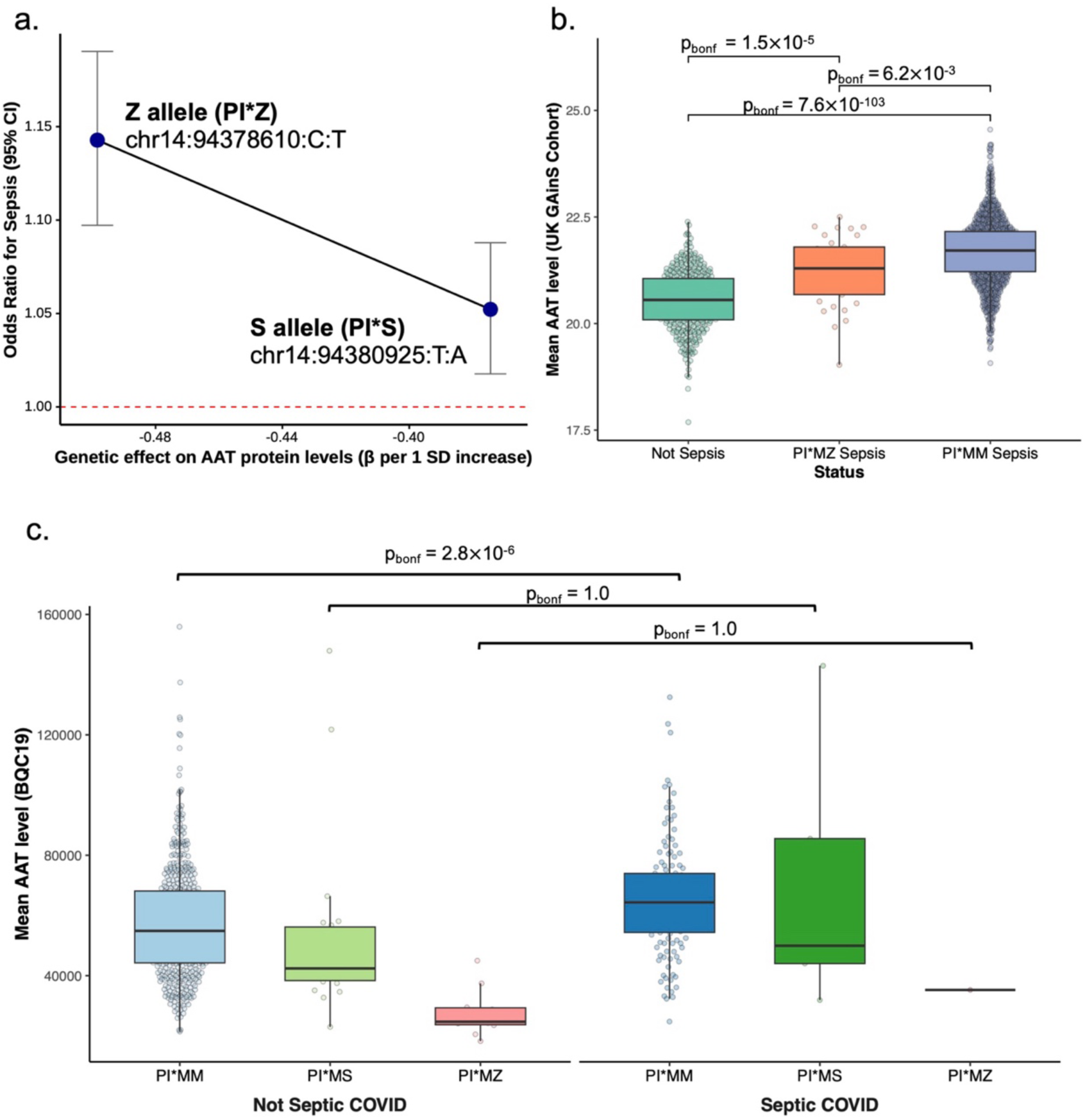
Allelic dose-response and dynamic proteomic validation of AAT levels during acute sepsis. a) Genetic dose-response relationship between SERPINA1 variants and sepsis risk. The x-axis represents the effect size (β) of the Z and S alleles on circulating AAT levels derived from UKB-PPP pQTL data. The y-axis indicates the corresponding risk of sepsis (odds ratio with 95% confidence intervals). The dashed red line denotes an OR of 1.0. b) Circulating AAT abundance in the UK GAinS cohort. Protein levels are compared across non-sepsis controls, sepsis patients with the wild-type genotype (PI*MM), and heterozygous Z-allele carriers with sepsis (PI*MZ). c) Circulating AAT abundance in the BQC19 cohort. Patients are stratified by sepsis status (non-septic vs. septic COVID-19) and *SERPINA1* genotype (PI*MM, PI*MS, and PI*MZ). For boxplots in b and c, the center line represents the median, box limits indicate the upper and lower quartiles (interquartile range). Each data point represents an individual participant. Statistical significance between groups was assessed using Wilcoxon rank-sum tests with Bonferroni correction (P_bonf_).

### Cross-phenotype MR and sensitivity analyses support independent protection

To explore the functional specificity of this protection, we performed a multi-trait MR analysis across several disease categories. Genetically predicted higher AAT levels were protective across a spectrum of infection-related phenotypes, pneumonia (OR = 0.90, P = 5.0 × 10^-8^; UKB-PPP), urinary tract infection (OR = 0.94, P = 0.049; UKB-PPP), skin infection (OR = 0.91, P = 7.7 × 10^-3^; UKB-PPP), bronchiectasis (OR = 0.58, P = 1.6 × 10^-21^; UKB-PPP) (which is thought to result from sequelae of past respiratory tract infections^42^) (Fig. 3). We also observed protective effects for positive control conditions highly relevant to sepsis progression and AAT biology, such as chronic obstructive pulmonary disease (COPD)^43^ (OR = 0.80, P = 1.9 × 10^-17^; UKB-PPP) and cirrhosis^35^ (OR = 0.81, P = 1.4 × 10^-11^; UKB-PPP). In contrast, AAT demonstrated opposite association with negative control of non-inflammatory metabolic conditions, such as type 2 diabetes (T2D) (OR = 1.09, P = 0.02; UKB-PPP) and hypertension (OR = 1.10, P = 1.3 × 10^-3^; UKB-PPP). Steiger tests were performed to ensure the correction causal direction (Supplementary Table 4). These results underscore that AAT’s protective role is not a universal effect but is specifically tuned to acute infection and lung/liver integrity, while potentially exerting distinct pleiotropic effects on metabolic risk.

Because *SERPINA1* variants are known to interact with environmental factors and contribute to chronic conditions that increase sepsis susceptibility, we conducted two separate sensitivity analyses in the UKB to isolate the direct genetic effect. First, to ensure the association was not driven by behavioral risk factors, we adjusted for smoking status; the *SERPINA1* missense variant remained significantly associated with sepsis risk (OR = 1.21, P = 6.6 × 10^-4^; Supplementary Table 5) and *SERPINA1* Z allele showed no interaction with smoking status. Second, we adjusted for a composite of clinical comorbidities (COPD, cirrhosis, and bronchiectasis before sepsis diagnosis) to determine if the effect was mediated by pre-existing organ damage. Even after accounting for these structural conditions, the association remained significant, (OR = 1.12, P = 5.7 × 10^-3^). Collectively, these results confirm that *SERPINA1* influences sepsis pathogenesis independently of smoking history or chronic structural disease, highlighting its role as a direct modulator of the systemic inflammatory response.

### Patient-level proteomic validation in UK GAinS cohorts

Our previous analyses used pQTLs derived under physiological conditions^40,41^, rather than during acute illness. To determine if AAT levels are dynamic during sepsis, we examined mass-spectrometry based proteomics from the UK GAinS cohort^21^. This study included 1,225 patients with pneumonia-related or abdominal sepsis and 372 non-sepsis controls (median age 65 years; 54% male). Participants were stratified by clinical phenotype and *SERPINA1* genotype into three groups: non-sepsis controls (n = 372), sepsis patients with wild-type alleles (PI*MM; n = 1,192), and sepsis patients heterozygous for the Z allele (PI*MZ; n = 33). Notably, genotype data were available exclusively for the sepsis cases. No Z-allele homozygotes (PI*ZZ) were identified in this cohort, and the S allele was excluded from the analysis due to a low genotyping call rate (< 20%).

During sepsis, AAT levels increased significantly in both genotype groups compared to non-sepsis controls (Wilcoxon rank-sum tests; P_bonf_ = 7.6 × 10^-103^ for PI*MM; P_bonf_ = 1.5 × 10^-5^ for PI*MZ). However, this acute-phase surge was significantly attenuated in PI*MZ carriers relative to the wild-type PI*MM group (P_bonf_ = 6.2 × 10^-3^; Fig 4b). These findings suggest that while individuals with genetically reduced AAT can mount an acute-phase response, their protective capacity is substantially blunted. This reinforces the hypothesis that the observed sepsis risk is driven by a relative functional deficiency during hyperinflammation, rather than the absolute absence of AAT characteristic of severe AATD (PI*ZZ).

### Patient-level proteomic validation in BQC19 cohorts

We replicated these temporal dynamics in the BQC19 cohort^22^, comprising patients with COVID-19-related sepsis and non-septic COVID-19 controls (median age 70 years; 55% male). Because genotyping was available for the entire cohort, participants were stratified into six subgroups based on sepsis status (septic COVID and not septic COVID) and *SERPINA1* genotype (PI*MM, PI*MS, and PI*MZ) (Fig. 4c). Consistent with our UK GAinS analysis, AAT levels were significantly elevated during septic COVID-19 in the wild-type PI*MM group (n = 133 septic vs. 504 non-septic; Wilcoxon rank-sum tests; P_bonf_ = 2.8 × 10^-6^). Although a similar upward trend was observed among PI*MZ and PI*MS carriers, the limited number of variant carriers (PI*MZ: 1 septic, 10 non-septic; PI*MS: 5 septic, 22 non-septic) precluded adequately powered statistical comparisons of the attenuated response.

To directly interrogate the protease-antiprotease balance, we analyzed the abundance of AAT alongside its primary target, ELANE, across the broader BQC19 general population. Participants were divided into three clinical states based on days since symptom onset (DSO) and disease severity: baseline convalescent (proteomics measured > 60 DSO; n = 168), not septic COVID (measured 0-60 days; n = 531), and septic COVID (measured 0-60 days; n = 139). We observed a stepwise increase in AAT abundance mirroring disease severity. Septic COVID-19 patients exhibited the highest AAT levels, significantly elevated compared to both the baseline group (Wilcoxon rank-sum tests; P_bonf_ = 6.7 × 10^-33^) and the not septic COVID group (P_bonf_ = 4.0 × 10^-6^). Not septic COVID patients also showed elevated AAT compared to baseline (P_bonf_ = 5.8 × 10^-34^) but remained lower than the septic cohort (Fig. 5a). Crucially, this overall rise in AAT was paralleled by a significant, concurrent increase in ELANE across the three groups. This parallel trajectory supports the hypothesis that AAT is acutely recruited to counteract excessive neutrophil elastase activity during severe infection.

**Figure 5.**
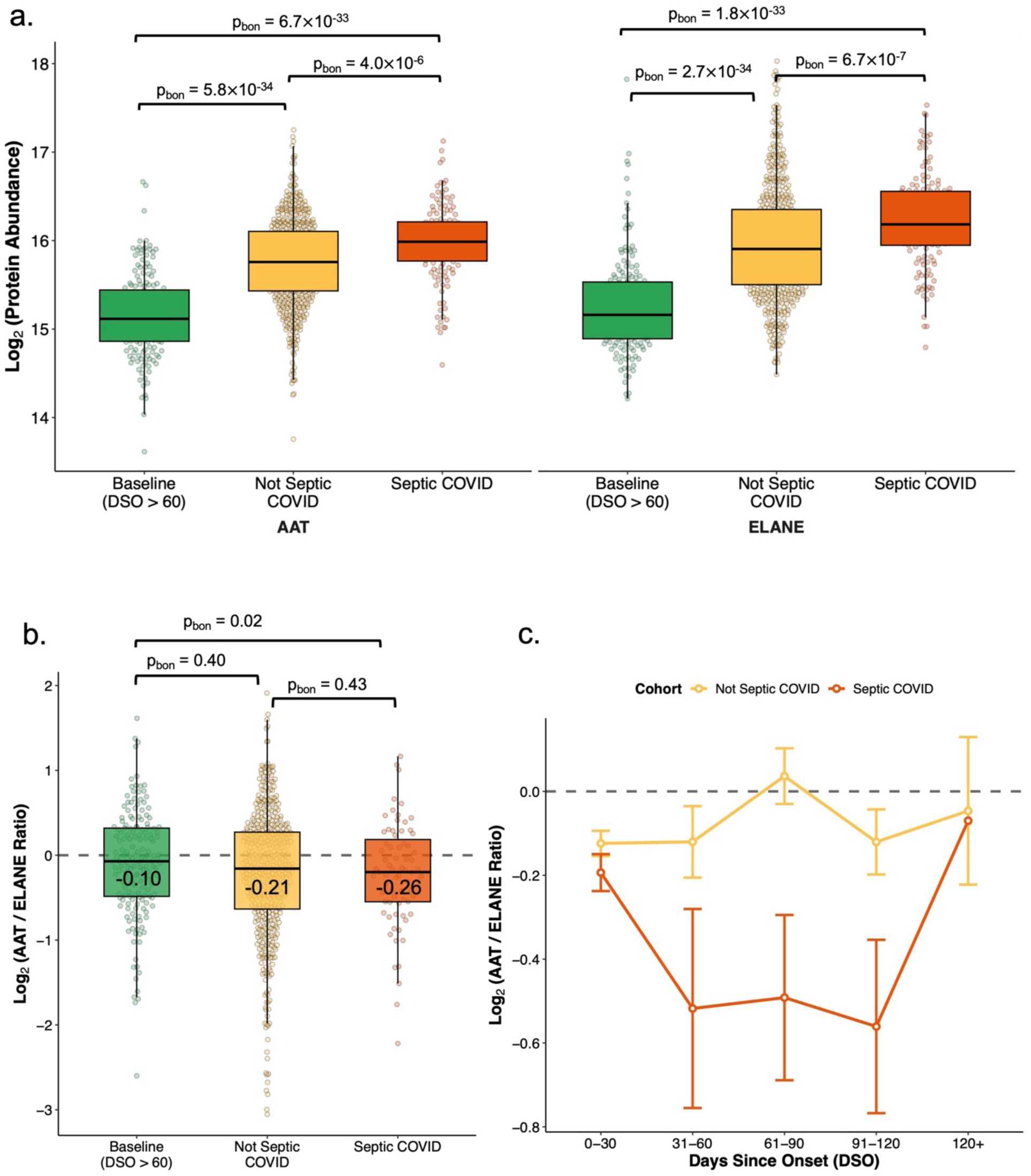
Dynamic proteomic validation of AAT and ELANE abundance and the protease-antiprotease balance during acute sepsis. a) log_2_-transformed circulating AAT and ELANE abundance in the BQC19 cohort. Patients are stratified by clinical status: baseline, non-septic COVID-19, and septic COVID-19. b) log_2_-transformed AAT/ELANE abundance ratio across the same clinical strata. The horizontal dotted line at y = 0 represents a 1:1 AAT/ELANE ratio. A lower ratio indicates a relative excess of ELANE, reflecting a disrupted protease-antiprotease balance. c) Longitudinal trajectory analysis of the AAT/ELANE ratio comparing non-septic and septic COVID-19 patients over time. Data points represent the mean log_2_(AAT/ELANE) ratio within each time bin. Error bars denote standard error of the mean. For boxplots in a and b, the center horizontal line represents the median, box limits indicate the upper and lower quartiles. Each underlying data point represents an individual participant. Statistical significance between groups was assessed using Wilcoxon rank-sum tests followed by Bonferroni correction (P_bonf_).

To quantify this dynamic, we calculated the log-transformed AAT/ELANE abundance ratio. In this scaled metric, a ratio of zero represents a perfect protease-antiprotease balance, while a negative ratio indicates a relative excess of ELANE, predisposing the host to protease-mediated damage. The baseline convalescent group maintained the highest, most balanced ratio (-0.10). In contrast, this balance was progressively disrupted during acute illness, dropping to-0.21 in non-septic COVID-19 (P_bonf_ = 0.40 vs. baseline) and declining significantly to-0.26 in septic COVID-19 (P_bonf_ = 0.02 vs. baseline) (Fig. 5b).

Longitudinal trajectory analyses further confirmed that the AAT/ELANE ratio remained consistently lower in septic patients compared to non-septic patients throughout the acute phase (Fig. 5c). To ensure these visual trajectories and statistical findings were not confounded by immunomodulatory treatments, we performed a sensitivity analysis adjusting for recent steroid administration (e.g., dexamethasone, hydrocortisone, prednisone) and interleukin-6 (IL-6) inhibitors (tocilizumab, sarilumab) before 5 days of the blood draw. After excluding these treated individuals (n = 6 baseline; n = 59 non-septic; n = 68 septic), the drug-adjusted results maintained the same pattern of protease-antiprotease mismatch (Supplementary Fig. 3, Supplementary Table 6).

Finally, AAT levels were significantly elevated during the first 30 days post-symptom onset in the septic COVID group (P = 4.4 × 10^-6^) but converged strictly with control levels following disease resolution (Supplementary Fig. 4). This return to baseline confirms that AAT acts as a highly specific acute-phase reactant during sepsis rather than being constitutively elevated.

Taken together, these data demonstrate that increasing disease severity drives a progressive disruption of the protease-antiprotease balance, suggesting that AAT supplementation during the acute phase could serve as a therapeutic intervention.

### AAT MR analyses are concordant with sepsis clinical trial results

To explore the downstream pathways influenced by AAT, we performed bidirectional MR testing AAT pQTLs against the circulating protein levels in the UKB-PPP. After Bonferroni correction (P < 1.0 × 10^-5^), 51 proteins were significantly associated with genetically predicted AAT levels (Supplementary Table 7). These proteins were categorized into five functional domains: coagulation/hemostasis, endothelial integrity, metabolic stress response, immune activation, and tissue damage/repair (Fig. 6).

**Figure 6.**
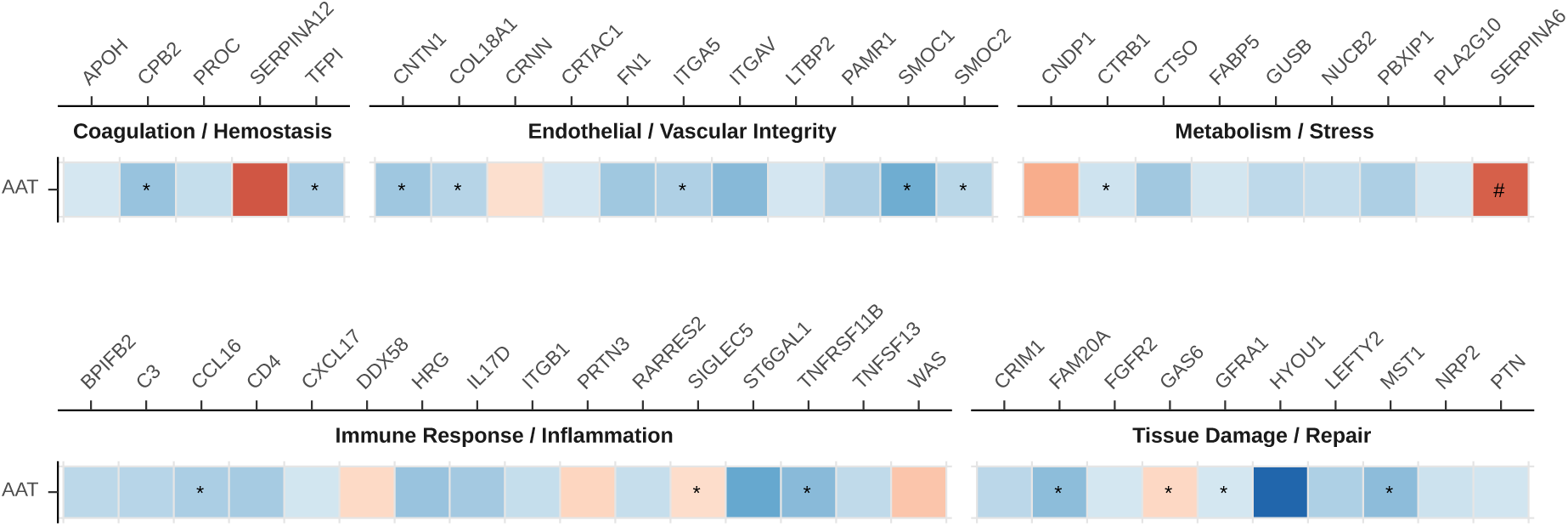
Mapping the AAT-regulated proteome and therapeutic pathways. Visualization of the circulating proteome associated with AAT as identified through proteome-wide Mendelian randomization. A total of 51 significant associations (P < 1.0 × 10^-5^) were identified and categorized into five functional domains: coagulation/hemostasis, endothelial/vascular integrity, metabolism/stress, immune response/inflammation, and tissue damage/repair. Red indicates positive effect sizes (positive correlation with AAT levels), while blue indicates negative effect sizes. AAT levels serve as the exposure for all displayed associations, except for those marked with a hash (#), which denote reverse MR results where AAT is the outcome. An asterisk (*) indicates associations that were independently replicated in the deCODE dataset with Bonferroni-corrected threshold. This proteogenomic map provides a genetic rationale for the diverse physiological roles of AAT and aligns with the observed outcomes of multiple historical sepsis clinical trials.

Several of these proteins have been the subject of previous randomized controlled trials (RCTs). For example, PROC (protein C)^44^ and TFPI (tissue factor pathway inhibitor)^45^ were tested for their anti-coagulation properties. However, these trials failed to show a reduction in mortality and were associated with an increased risk of bleeding^9,10,46,47^. Our MR results are concordant with these findings: higher AAT genetically reduces PROC and TFPI, which is the opposite of the intervention tested in their respective trials.

Furthermore, we identified a reciprocal genetic relationship between AAT and SERPINA6, the primary cortisol-binding globulin^48^. RCTs show that corticosteroids remain one of the few pharmacological interventions shown to reduce mortality in septic shock^49^, the reciprocal increases between these two serpins provide a plausible mechanistic link between AAT and the stabilization of the host’s endocrine response to stress.

Other associated proteins, including HYOU1^50^, FN1^51^, SMOC1, and SMOC2^52,53^, have been linked to fibrotic pathways. The reduction of these proteins by AAT suggests a potential role for AAT in mitigating multi-organ fibrosis, potentially by inhibiting the formation of neutrophil extracellular traps (NETs)^54^.

### Expansion to Diverse Genetic Ancestries

Recognizing the importance of genetic diversity in locus discovery, we conducted ancestry-specific genome-wide association studies (GWAS) of sepsis in African (7,366 cases; 167,270 controls) and Admixed American (6,088 cases; 103,762 controls) populations. Additionally, we extracted summary statistics for East Asian (2,022 cases; 339,066 controls) and South Asian (1,146 cases; 47,343 controls) cohorts from publicly available sources (Supplementary Table 1, Supplementary Fig. 5-8). As previously noted, the pathogenic SERPINA1 Z allele is virtually absent in these non-European cohorts, underscoring the need to identify ancestry-specific risk loci.

In the African ancestry analysis, we identified one genome-wide significant locus (P < 5 ×10^-8^) at chr5:161537792:C:T (OR = 1.15, 95% CI = 1.10-1.21, P = 1.7 × 10^-8^) (Supplementary Table 8). This variant maps closely to GABRB2, which encodes a GABA receptor subunit. Recent studies have demonstrated that epilepsy syndromes associated with GABRB2 variants are highly sensitive to fever and systemic infections^55^, suggesting a potential neuro-immune intersection during illness.

In the Admixed American cohort, a single intronic variant reached genome-wide significance (chr18:8443061:TG:T; OR = 0.90, 95% CI = 0.86-0.93, P = 1.2 × 10^-8^). This represents a novel locus that has not previously been associated with sepsis-related phenotypes. Finally, no variants reached genome-wide significance in the East Asian or South Asian analyses; this is likely due to smaller sample sizes limiting statistical power in these strata.

## Discussion

This study integrates evidence across large-scale human genetics, causal inference, and patient proteomics to demonstrate that *SERPINA1* and its encoded protein, AAT, play a central role in sepsis susceptibility and host response. AAT is a canonical acute-phase reactant and is normally upregulated during sepsis, such that circulating levels rise in response to infection. Our meta-analysis of over 60,000 sepsis cases identified the pathogenic Z allele as a primary risk locus, a variant long known to impair AAT folding and secretion.

MR and colocalization results provide robust causal evidence for this relationship. Genetically predicted higher AAT levels were protective across a broad spectrum of infectious phenotypes, such as pneumonia, urinary tract infections, and skin infections. While it is unlikely that AAT plays a role in each of these infections, we here used the fact that genetic biobanks are biased towards more critical illness that require hospital admissions. Hence, our results likely reflect that AAT is protective for severe infection presentations in general, which is consistent with being protective for sepsis. This functional specificity, combined with the allelic dose-response observed between the severe Z and milder S alleles, indicates that circulating AAT levels directly dictate the severity of the inflammatory cascade. Clinical proteomic validation in the UK GAinS and BQC19 cohorts further refined this model. We observed that while heterozygous Z-allele carriers (PI*MZ) still mount a significant acute-phase AAT surge compared to controls during sepsis, this response is severely blunted relative to wild-type (PI*MM) individuals. This demonstrates that the genetic mutation creates a secretion bottleneck during extreme physiological demand, preventing these patients from reaching fully protective AAT thresholds. Consequently, their increased sepsis risk stems from a relative functional deficiency rather than an absolute absence of the protein. Notably, by measuring AAT alongside its primary target, ELANE, we demonstrated that the profound surge in neutrophil elastase outpaces the host’s maximum AAT biosynthetic capacity during extreme hyperinflammation, creating a vulnerability to protease-mediated damage even in wild-type individuals. This shifts the paradigm of AAT from a targeted treatment for rare genetic deficiencies to a broadly applicable adjunctive therapy for severe sepsis, regardless of underlying genotype or genetic ancestry.

Crucially, the AAT is already pharmacologically tractable. Intravenous AAT augmentation therapy is currently approved for AAT-deficient lung disease, and our genetic and proteomic results strongly suggest it could be repurposed as an acute intervention for sepsis. This hypothesis is supported by independent computational network analyses using gene expression profiles^39^, as well as experimental mouse models demonstrating improved survival following AAT administration^56^. Furthermore, functional studies of the AAT Pittsburgh mutation, which fatally alters the enzyme’s protease target specificity, demonstrated deleterious sepsis outcomes in primate models^57,58^, underscoring the protective role of normal endogenous AAT activity during systemic inflammation.

While other pharmacological approaches exist, they are suboptimal for acute sepsis. For example, the dipeptidyl peptidase 1 (DPP1) inhibitor Brensocatib successfully targets neutrophil serine proteases (including ELANE) in bronchiectasis^59^; however, its mechanism relies on the gradual depletion of proteases during neutrophil maturation, making its prolonged clinical course poorly suited to combat a rapid-onset septic protease storm. Alternatively, Ulinastatin, a broad-spectrum serine protease inhibitor, is clinically utilized for acute circulatory failure and sepsis in China. However, to our knowledge, this has never been tested in a robust pre-specified large-scale high-quality randomized trial^60^.

Another strength of this study is the integration of our findings with existing clinical trial data. Our genetic evidence aligns with the result of PROC^9^ and TFPI^10^ trials, which did not show clinical benefit, while increasing severe bleeding complications. Conversely, the reciprocal relationship between AAT and the corticosteroid-binding protein SERPINA6 provides a genetic echo of the mortality benefits observed with steroid administration in septic shock^49^. This provides orthogonal evidence that our results are robust and more likely to translate well to AAT supplementation trial.

Several limitations warrant consideration. First, while we demonstrated a clear systemic effect, the exact timing and dosage for clinical intervention remain to be determined. Second, although our analysis included both bacterial and viral sepsis (COVID-19), it remains unclear which specific pathogens are most influenced by AAT activity. Future studies with more granular microbiological data will be required to determine if AAT protection is pathogen-agnostic or varies between infectious etiologies. Third, although the signal is not exclusively driven by homozygous carriers, the degree to which AAT influences specific organ failures, such as respiratory distress versus circulatory shock, requires further granularity. Finally, our proteomic validation was limited to circulating levels; tissue-specific dynamics, particularly in the liver and lung, remain to be explored.

In conclusion, these findings position AAT as a biologically coherent and genetically validated candidate for sepsis therapeutics. While previous experimental and observational studies have suggested the therapeutic potential of AAT, this work provides the first large-scale human genetic evidence linking AAT directly to sepsis susceptibility and outcomes. Given that drug targets supported by human genetics are significantly more likely to achieve success in clinical development^18^, our results provide a strong evidence-based foundation for future trials.

## Methods

### Sepsis GWAS meta-analysis

We conducted a fixed effects meta-analysis of sepsis in METAL^23^ across four large-scale biobanks: UKB^24^, MVP^25^, FinnGen(R12)^26^ and AoU^27^. The total sample size included 60,314 sepsis cases and 1,464,733 controls (Supplementary Table 1). Summary statistics for UKB and AoU were generated using REGENIE^61^ to account for population stratification and cryptic relatedness, while summary statistics for MVP and FinnGen were obtained from their respective public repositories. To identify independent signals, we utilized PLINK v2.0 with the UKB European reference panel to clump (r^2^ threshold = 0.001) genome-wide significant SNPs (P < 5 × 10^-8^) within a 1000 kb distance^62,63^.

### Cohort descriptions and phenotyping

#### UK Biobank

The UK Biobank is a prospective population-based study comprising approximately 500,000 participants from the United Kingdom, characterized by extensive phenotypic and genotypic data^24^. To define a comprehensive sepsis cohort, identified cases via the first occurrence of streptococcal septicaemia (field 130068, ICD-10 code A40), other septicaemia (field 130070, ICD-10 code A41), and puerperal sepsis (field 132286, ICD-10 code O85). Following these definitions, we conducted a GWAS among individuals of European ancestry with 17,569 cases and 444,032 controls. To account for population structure and cryptic relatedness, association testing was performed using REGENIE with covariates of age, sex, age^2^, the first 10 genetics principal components (PCs), enrolment enters, and genotype array.

#### All of Us (AoU)

The NIH All of Us Research Program is a nationwide longitudinal initiative established to catalyze precision medicine and address historical imbalances in genomic science^27^. Our study leveraged the version 8 data release, which provides clinical-grade whole-genome sequencing for a cohort of over 414,000 participants. We retained individuals with DNA samples that passed quality control and with available electronic health records.

Cases of sepsis were defined based on standard concept identifiers (concept ID 132797) from the Observational Medical Outcomes Partnership (OMOP) Common Data Model provided by the AoU. Ancestry-specific GWAS were performed using REGENIE, adjusting for age, age^2^, sex, and the first 10 genetic principal components, across European (8,791 cases; 162,220 controls), African (2,930 cases; 53,865 controls), and Admixed American (4,087 cases; 47,708 controls) ancestry populations.

Cases of bronchiectasis were defined based on concept ID 256449 (Supplementary Table 9). Cases of pneumonia were defined based on concept ID 255848. Cases of COPD were defined based on concept ID 255573.

#### Million Veterans Program (MVP)

We used publicly available GWAS summary statistics from the MVP^25^. We selected the case-control GWAS of sepsis using PheCode 994 in European (16,821 cases; 419,433 controls), African (4,436 cases; 113,405 controls) and Admixed American (2,001 cases; 56,054 controls) populations.

To evaluate related respiratory and systemic conditions, we additionally retrieved European-ancestry GWAS data for the following traits: bronchiectasis (PheCode 496.3; 1,877 cases, 447,375 controls), pneumonia (PheCode 480; 43,748 cases, 378,820 controls), chronic obstructive pulmonary disease (COPD; PheCode 496.21; 24,495 cases, 409,429 controls), and hypertension (PheCode 401; 322,448 cases, 105,690 controls).

#### FinnGen

We used publicly accessible GWAS summary statistics from the FinnGen Release 12, a nationwide biobank linking genomic data with digital healthcare recores from the Finnish population^26^. For our primary analysis, we obtained data for the “other sepsis” phenotype (FinnGen endpoint: AB1_OTHER_SEPSIS; ICD-10: A41), which comprised 17,113 cases and 439,048 controls. Because FinnGen does not provide a pre-computed GWAS for a composite “all sepsis” trait, and individual-level data were unavailable for custom aggregation, we selected “other sepsis” as it represents the largest available sepsis case cohort in the dataset.

Additionally, we extracted GWAS summary statistics for related respiratory conditions, including bronchiectasis (endpoint: J10_BRONCHIECTASIS; 2,932 cases, 409,070 controls), pneumonia (endpoint: J10_PNEUMONIA; 78,791 cases, 421,557 controls), and chronic obstructive pulmonary disease (COPD; endpoint: J10_COPD; 24,138 cases, 409,070 controls).

#### Genes & Health study

We utilized publicly available GWAS summary statistics from the Genes & Health study (www.genesandhealth.org), a long-term cohort comprising approximately 100,000 individuals of South Asian (primarily Bangladeshi and Pakistani) ancestry. Specifically, we extracted data for the sepsis phenotype defined by ICD-10 code A41. Similar to our approach with the FinnGen dataset, because a pre-computed composite “all sepsis” trait is unavailable and we lack access to individual-level data for custom phenotyping, we extracted A41 to represent sepsis in this cohort (1,146 cases; 47,343 controls).

#### Taiwan Precision Medicine Initiative

To expand our analysis into East Asian populations, we utilized GWAS summary statistics from the Taiwan Precision Medicine Initiative (TPMI) dataset^64^. This nationwide consortium features linked genomic and clinical data for 565,390 participants. Summary statistics for the “Sepsis and SIRS” phenotype (PheCode: 994) were downloaded directly from the TPMI PheWeb (https://pheweb.ibms.sinica.edu.tw/).

### Mortality analyses

We performed a retrospective cohort study within the UKB to evaluate the clinical impact of the identified risk alleles on sepsis outcomes. The primary endpoint was 30-day sepsis mortality, defined as death occurring within a window spanning five days prior to and 30 days following the initial sepsis diagnosis (streptococcal septicaemia (field 130068), other septicaemia (field 130070), and puerperal sepsis (field 132286)). Genotype dosages for the lead variants were extracted and computed using PLINK v2.0. We employed logistic regression, adjusted for age and sex, to estimate the odds of 30-day mortality associated with each risk allele compared to the general UKB population (3,131 cases; 457,750 controls). To distinguish between genetic drivers of disease susceptibility and disease severity, we conducted a secondary subgroup analysis restricted to the sepsis cohort, comparing sepsis-related fatalities (n = 3,131) directly against sepsis survivors (n = 14,334).

### Two-sample Mendelian randomization

We conducted two-sample Mendelian randomization (MR) to evaluate the causal relationship between circulating protein levels and diverse clinical outcomes using the TwoSampleMR R package.

### Genetic instruments and pQTL data

Genetic instruments for AAT were two missense variants in *SERPINA1* (chr14:94378610:C:T and chr14:94380925:T:A) pQTLs in both UKB-PPP^41^ (Olink platform; 34,557 European individuals) and deCODE^40^(SomaScan platform; 35,559 individuals). The F-statistics for UKB-PPP genetic instruments are 345 for Z allele and 451 for S allele. The F-statistics for deCODE genetic instruments are 2973 and 2680 respectively. *Cis*-pQTLs were defined as variants located within ±500 kb of the gene’s transcription start site to capture proximal regulatory and functional variation.

### Outcome phenotypes

Clinical outcomes included sepsis, bronchiectasis, pneumonia, chronic obstructive pulmonary disease (COPD), urinary tract infection (UTI), and bacterial skin infections. Metabolic traits (type 2 diabetes (T2D) and hypertension) and cirrhosis served as control or secondary phenotypes.

Summary statistics for cirrhosis^65^, T2D^66^, and hypertension^25^ were obtained from established public repositories.

UTI and bacterial skin infection GWAS were generated specifically within the UKB using REGENIE based on ICD-10 codes (UTI: N10, N30, N39; bacterial skin infection: L01-04, L8 and A46) (Supplementary Table 10).

Sepsis, pneumonia and bronchiectasis results were derived from a fixed-effects meta-analysis (METAL) of the UKB, MVP, FinnGen, and AoU cohorts (Supplementary Table 9).

The MR analysis across phenotypes utilized the Inverse-Variance Weighted (IVW) method with both fixed and random effects. To account for multiple testing, significance was determined using a Bonferroni corrected P value threshold (P < 0.005 [0.05/9 phenotypes]).

### Protein-protein bidirectional MR

To identify downstream pathways regulated by AAT, we extended our MR framework to examine the association between AAT pQTLs and the broader circulating proteome (2,923 proteins in UKB-PPP). In this discovery phase, AAT served as the exposure and other plasma proteins as outcomes. We subsequently performed reverse MR, where other proteins were treated as exposures and AAT as the outcome, to identify potential upstream regulators or feedback loops. Instrument selection and procedure followed the original MR analyses. All associations were assessed for significance using Bonferroni correction for the total number of protein pairs tested. Validation MR was performed using deCODE data. To account for multiple testing, we used Bonferroni-corrected P value threshold (P < 1.0 × 10^-5^ [0.05/ (2,939 AAT on other proteins + 1,936 other proteins on AAT)]).

### Colocalization of European ancestry meta-analyzed GWAS with pQTLs

To investigate whether the genetic signals for sepsis and circulating AAT concentrations share a common causal variant, we performed pairwise conditional colocalization using PWCoCo with UKB European reference panel^67^. This method extends the traditional colocalization framework by utilizing conditional summary statistics, thereby accounting for potential multiple independent signals within the *SERPINA1* locus. Analysis was conducted using default priors, with high confidence colocalization defined as a posterior probability PP.share ≥ 0.8.

## Sensitivity analyses

To determine if the association between *SERPINA1* and sepsis was independent of known alpha-1 antitrypsin deficiency-related comorbidities and environmental factors, we conducted two distinct logistic regression sensitivity analyses in the UK Biobank. In the first model, we adjusted for smoking status, categorized as never, former, or current smoker, to ensure the genetic signal was not primarily driven by behavioral or environmental risk factors. In the second model, we adjusted for a composite of clinical comorbidities including COPD, cirrhosis, and bronchiectasis occur before the earliest diagnosis of sepsis, to determine if the effect was mediated by preexisting structural organ damage. Both models were adjusted for age and sex, with results reported as odds ratios and 95% confidence intervals to isolate the direct effect of the *SERPINA1* genotype on sepsis pathogenesis.

### UK Genomic Advances in Sepsis (UK GAinS) cohort

To validate the influence of the AAT levels on the acute-phase response, we leveraged proteomic data from the UK GAinS cohort^21^, a discovery effort within the Genomic Advances in Sepsis (GAin) study. This cohort included individuals admitted to intensive care units with sepsis secondary to community-acquired pneumonia or fecal peritonitis, as well as non-sepsis controls. Plasma protein abundance was quantified using a high-throughput, liquid chromatography-mass spectrometry pipeline, providing a comprehensive map of the sepsis proteome.

Our analysis focused on a subset of 1,225 sepsis patients and 372 non-sepsis controls stratified by *SERPINA1* genotype. Participants were categorized into three groups: non-sepsis controls (n = 372), sepsis patients homozygous for the common allele (PI*MM, n = 1,192), and heterozygous Z-allele carriers with sepsis (PI*MZ; n = 33). The S-allele was excluded from the UK GAinS analysis due to a call rate of less than 20%. To assess differences in the magnitude of the protein response and we took mean AAT values when there are multiple measurements and compared using the Wilcoxon rank-sum test. All resulting P values were adjusted using the Bonferroni correction method to account for multiple testing. Protein measurement quality control and processing was described by Mi et al^21^.

### The Biobanque Québécois sur la COVID-19 (BQC19) cohort

Further validation was conducted using the BQC19, a province-wide initiative in Quebec designed to prospectively capture the clinical and biological determinants of COVID-19 trajectories^22^. The BQC19 recruits both hospital-based participants (with severe or critical illness) and community-based participants (with mild or asymptomatic infection) across multiple sites. For this analysis, severe COVID-19-related sepsis was defined by the clinical requirement for organ support (mechanical ventilation or vasopressors) in an intensive care setting^68^. Plasma concentrations of AAT and ELANE were quantified using SomaScan technology and reported in relative fluorescence units (RFU). Genotypes for each participant were extracted and processed using PLINK 2.0.

To standardize comparisons of AAT and ELANE protein abundances, as well as their ratio, all values were log_2_-transformed. To characterize the temporal dynamics of the acute-phase response, we modeled the longitudinal trajectory of AAT levels in the BQC19 cohort relative to the days since symptom onset (DSO). Given that participants provided multiple plasma samples over the course of their illness and recovery, we categorized measurements into discrete temporal windows: 0-30, 31-60, 61-90, 91-120, and >120 days post-onset. For each time bin, we calculated the mean AAT level (± standard deviation) and the mean AAT/ELANE ratio (± standard error) to visualize the evolution of the host response from the acute inflammatory stage through disease resolution. This binning approach allowed us to assess the temporal dynamics of AAT expression and the maintenance of AAT-ELANE homeostasis over time.

To prevent pharmacological confounding of the proteomic data, we excluded individuals who received systemic corticosteroids (dexamethasone, methylprednisolone, betamethasone, hydrocortisone, or prednisone) or IL-6 receptor antagonists (tocilizumab or sarilumab) within five days prior to blood collection. This ensured the observed protein abundances were not driven by drug-induced alterations.

### Ancestry-Specific Meta-Analyses

To evaluate genetic associations across diverse populations, we conducted ancestry-specific analyses for four additional major ancestry groups: African, Admixed American, East Asian, and South Asian (Supplementary Table 1). The meta-analyses were performed using a fixed-effects model implemented in the METAL for African and Admixed American populations. Summary statistics for the East Asian and South Asian cohorts were extracted from the previously mentioned repositories. Then, independent genome-wide significant SNPs were identified using the clumping in PLINK 2.0.

### Data Harmonization and Quality Control

To ensure cross-cohort comparability and minimize technical artifacts, all raw summary statistics underwent a standardized quality control pipeline. Variants were filtered to retain only those with a minor allele frequency (MAF) between 0.005 and 0.995, thereby mitigating bias from low-frequency variants with potentially poor imputation quality. We standardized variant nomenclature to a consistent “chromosome:position:ref:alt” format and harmonized all effect sizes and allele frequencies to the designated alternative allele to ensure directional consistency.

Data quality was further strictly controlled by filtering for imputation accuracy; variants were restricted to those with an R^2^ > 0.6. This stringent threshold was applied to eight of the ten datasets; however, the Genes & Health and TPMI cohorts were included without R^2^ filtering as these specific metrics were not available in their public summary statistics. Following these harmonization steps, the processed datasets were integrated for final downstream causal inference and meta-analysis.

## Data Availability

MVP, FinnGen, Genes & Health, and TPMI GWASs are publicly available on their respective websites. Accessing AoU and UKB data for GWAS requires an application through the respective official processes. We will upload the meta-analysis GWAS conducted in this study to GWAS Catalog upon publication.

Proteomics data of UK GAinS are available on the Proteomics Identification Database (PRIDE) (accession ID PXD039875). GAinS genotyping data were deposited at the European Genome-phenome Archive (EGA), under accession number EGAD00001015369^69^ and are available upon request. Proteomics and genomics data of BQC19 are available upon request (https://www.bqc19.ca/en/home/).

## Code Availability

We used R v.4.1.2,

PLINK v.2.0 (https://www.cog-genomics.org/plink/2.0/),

TwoSampleMR v. 0.6.4,

PWCoCo (https://github.com/jwr-git/pwcoco),

REGENIE v.4.1 (https://github.com/rgcgithub/regenie),

METAL (https://github.com/statgen/METAL),

LocusZoom v0.3.8

Custom code and scripts used to reproduce the analyses for this study will be made publicly available on GitHub (https://github.com/Dandan-Alina-Tan/sepsis_AAT) upon publication.

---

**Extended Data Figure 1.**
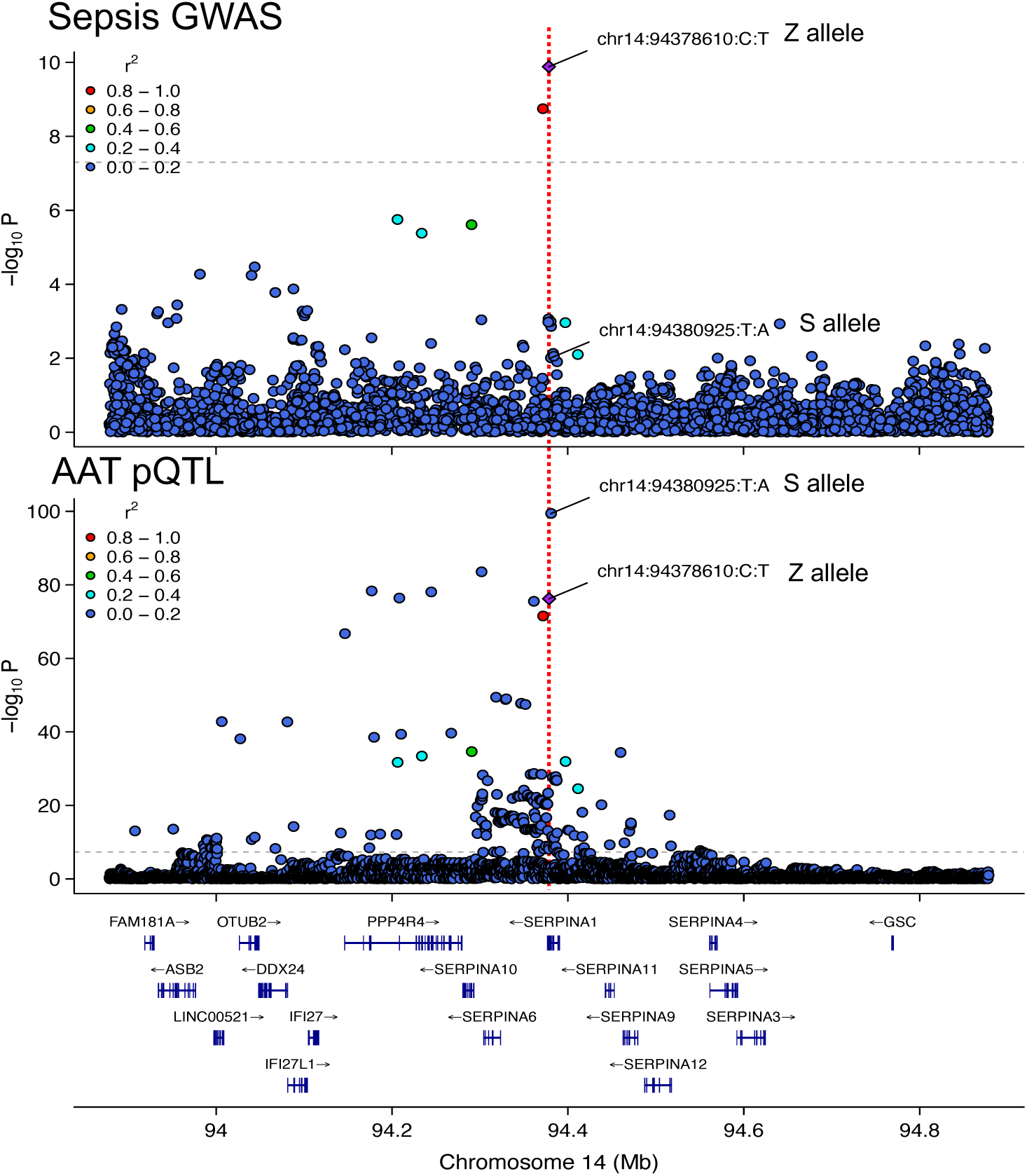
Colocalization of the sepsis GWAS signal and AAT pQTL at the SERPINA1 locus. Regional association (LocusZoom) plots comparing the sepsis meta-analysis (top) and circulating AAT protein quantitative trait loci (pQTL) from the UKB-PPP (bottom) on chromosome 14. The variants corresponding to the SERPINA1 Z allele (chr14:94378610:C:T, purple diamond) and S allele (chr14:94380925:T:A) are highlighted, with surrounding points colored by their linkage disequilibrium (r^2^) with the index variant. The high degree of regional similarity and a strong posterior probability (PP.share = 99%) from pairwise conditional colocalization indicate a shared causal genetic signal driving both AAT levels and sepsis risk.

## Acknowledgements

We gratefully acknowledge the participants and volunteers of the All of Us Research Program, Genes & Health, the Quebec COVID-19 Biobank (BQC19), and the UK Genomic Advances in Sepsis (GAinS) study; without their essential contributions, this research would not have been possible.

We thank the National Institutes of Health for providing access to the *All of Us* data. The BQC19 cohort is supported by the Fonds de recherche du Québec-Santé (FRQS), Génome Québec, the Public Health Agency of Canada, and the Ministère de la Santé et des Services sociaux. The UK GAinS study was supported by the Chinese Academy of Medical Sciences (CAMS) Innovation Fund for Medical Science (CIFMS), China (grant number: 2024-I2M-2-001-1). We thank Professor Vincent Mooser (McGill University) for his insightful discussions and valuable advice.

Finally, Google Gemini was used to refine the grammar and improve the writing style for clarity and conciseness.

## Conflict of Interest

J.B.R. acknowledges investigator-initiated grant funding to his institution from Roche, Eli Lilly, GlaxoSmithKline, and Biogen for projects unrelated to the submitted work. J.B.R. is the Chief Executive Officer of and holds equity in 5Prime Sciences (www.5primesciences.com), a company providing genetics-based drug development research services. K.Y.H.L. is an employee of 5Prime Sciences, and T.L. provides consulting services to the company. 5Prime Sciences was not involved in the design, execution, analysis, interpretation, or funding of this study, and these commercial relationships are unrelated to the current research. All other authors declare no competing interests.

### Gene & Health Funding Statement

Genes & Health is/has recently been core-funded by Wellcome (WT102627, WT210561), the Medical Research Council (UK) (M009017, MR/X009777/1, MR/X009920/1), Higher Education Funding Council for England Catalyst, Barts Charity (845/1796), Health Data Research UK (for London substantive site), and research delivery support from the NHS National Institute for Health Research Clinical Research Network (North Thames). We acknowledge the support of the National Institute for Health and Care Research Barts Biomedical Research Centre (NIHR203330); a delivery partnership of Barts Health NHS Trust, Queen Mary University of London, St George’s University Hospitals NHS Foundation Trust and St George’s University of London.

Genes & Health is/has recently been funded by Alnylam Pharmaceuticals, Genomics PLC; and a Life Sciences Industry Consortium of AstraZeneca PLC, Bristol-Myers Squibb Company, GlaxoSmithKline Research and Development Limited, Maze Therapeutics Inc, Merck Sharp & Dohme LLC, Novo Nordisk A/S, Pfizer Inc, Takeda Development Centre Americas Inc.

We thank Social Action for Health, Centre of The Cell, members of our Community Advisory Group, and staff who have recruited and collected data from volunteers. We thank the NIHR National Biosample Centre (UK Biocentre), the Social Genetic & Developmental Psychiatry Centre (King’s College London), Wellcome Sanger Institute, and Broad Institute for sample processing, genotyping, sequencing and variant annotation. This work uses data provided by patients and collected by the NHS as part of their care and support. This research utilised Queen Mary University of London’s Apocrita HPC facility, supported by QMUL Research-IT, http://doi.org/10.5281/zenodo.438045

We thank: Barts Health NHS Trust, NHS Clinical Commissioning Groups (City and Hackney, Waltham Forest, Tower Hamlets, Newham, Redbridge, Havering, Barking and Dagenham), East London NHS Foundation Trust, Bradford Teaching Hospitals NHS Foundation Trust, Public Health England (especially David Wyllie), Discovery Data Service/Endeavour Health Charitable Trust (especially David Stables), Voror Health Technologies Ltd (especially Sophie Don), NHS England (for what was NHS Digital) - for GDPR-compliant data sharing backed by individual written informed consent.

