## Supplemental Figures for "Human Genetic Analysis Reveals Circulating Alpha-1 Antitrypsin Level as a Protective Factor in Sepsis"

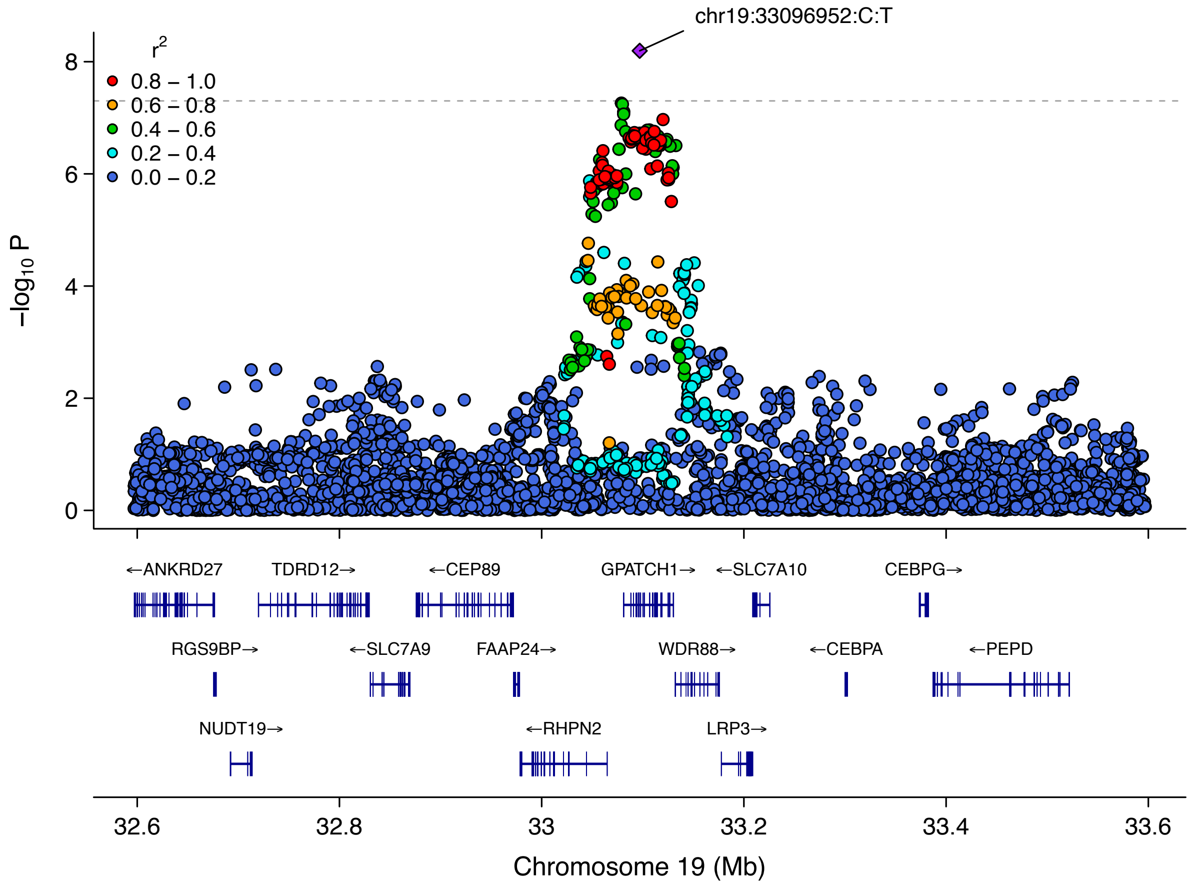


**Supplementary Figure 1. LocusZoom plot around the lead variant chr19:33096952:C:T (±500 kb)**

Individual data points denote genetic variants, color-coded by their degree of linkage disequilibrium ($r^2$) with the index variant based on a European reference panel. Genomic coordinates are plotted along the x-axis, while the y-axis represents the $-\log_{10}(P)$ significance levels derived from the sepsis association study. The dashed horizontal line marks the standard threshold for genome-wide significance ($P = 5 \times 10^{-8}$).

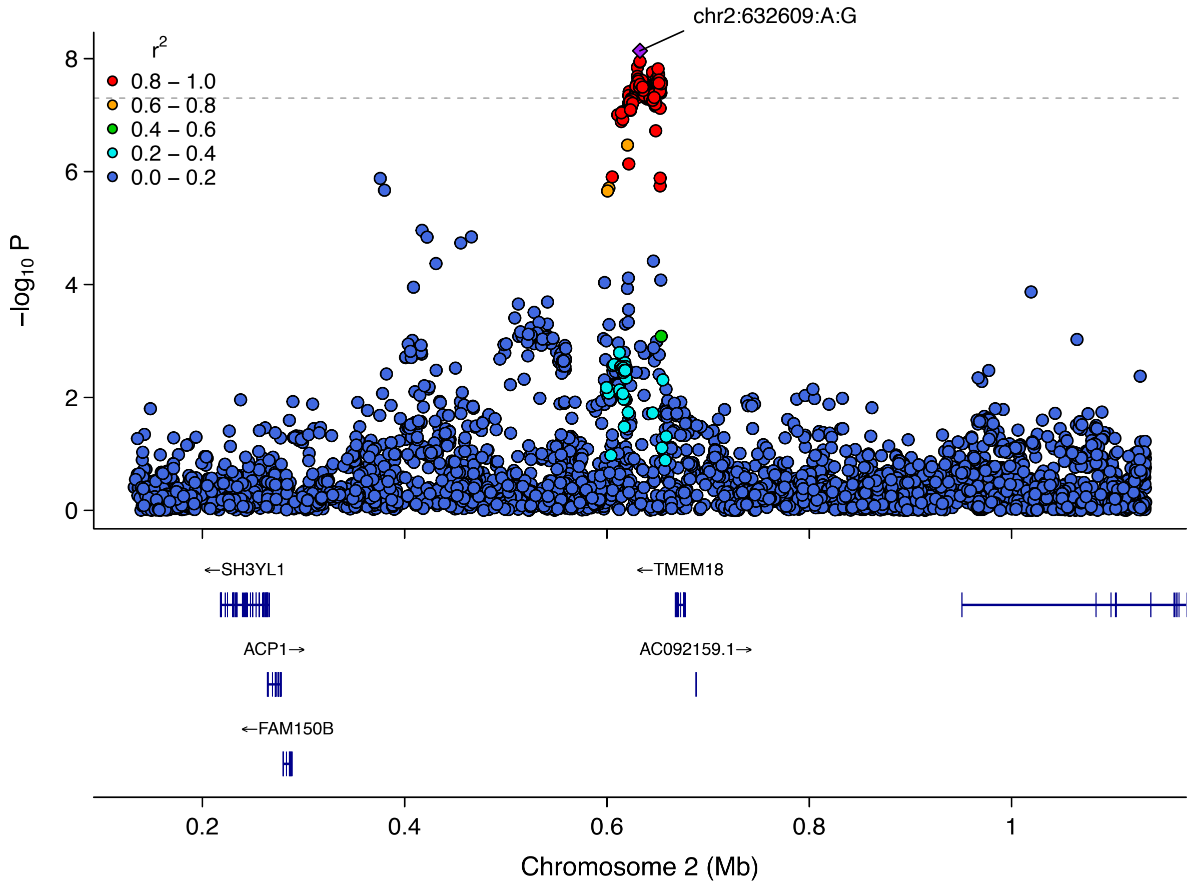


**Supplementary Figure 2. LocusZoom plot around the lead variant chr2:632609:A:G (±500 kb)**

Individual data points denote genetic variants, color-coded by their degree of linkage disequilibrium ($r^2$) with the index variant based on a European reference panel. Genomic coordinates are plotted along the x-axis, while the y-axis represents the $-\log_{10}(P)$ significance levels derived from the sepsis association study. The dashed horizontal line marks the standard threshold for genome-wide significance ($P = 5 \times 10^{-8}$).

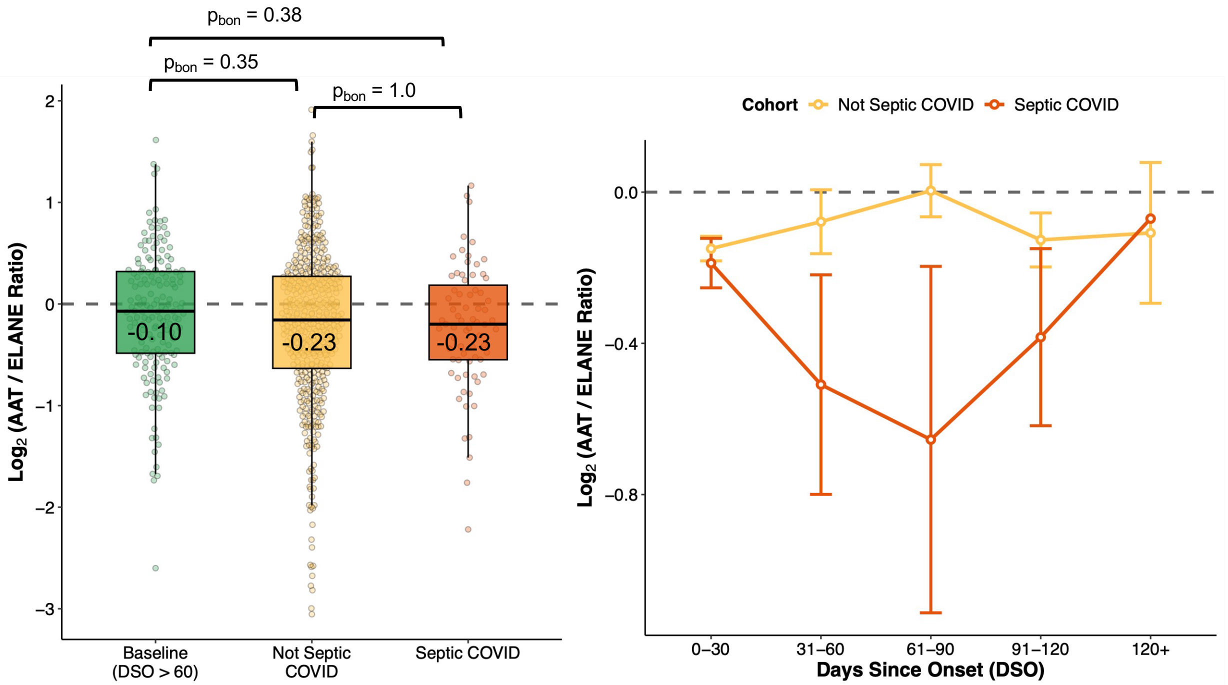


**Supplementary Figure 3. Drug-adjusted analysis of AAT and ELANE dynamics in the BQC19 cohort.** All panels reflect data following adjustment for recent immunomodulatory treatments. Patients are stratified by clinical status: baseline convalescent, non-septic COVID-19, and septic COVID-19. **a)** $log_2$-transformed circulating AAT and ELANE abundance. **b)** The corresponding $log_2$(AAT/ELANE) abundance ratio across the septic COIVD and not septic COVID. Consistent with the primary analysis, a lower ratio indicates a relative excess of ELANE and a disrupted protease-antiprotease balance. Statistical significance between groups was assessed using Wilcoxon rank-sum tests followed by Bonferroni correction.

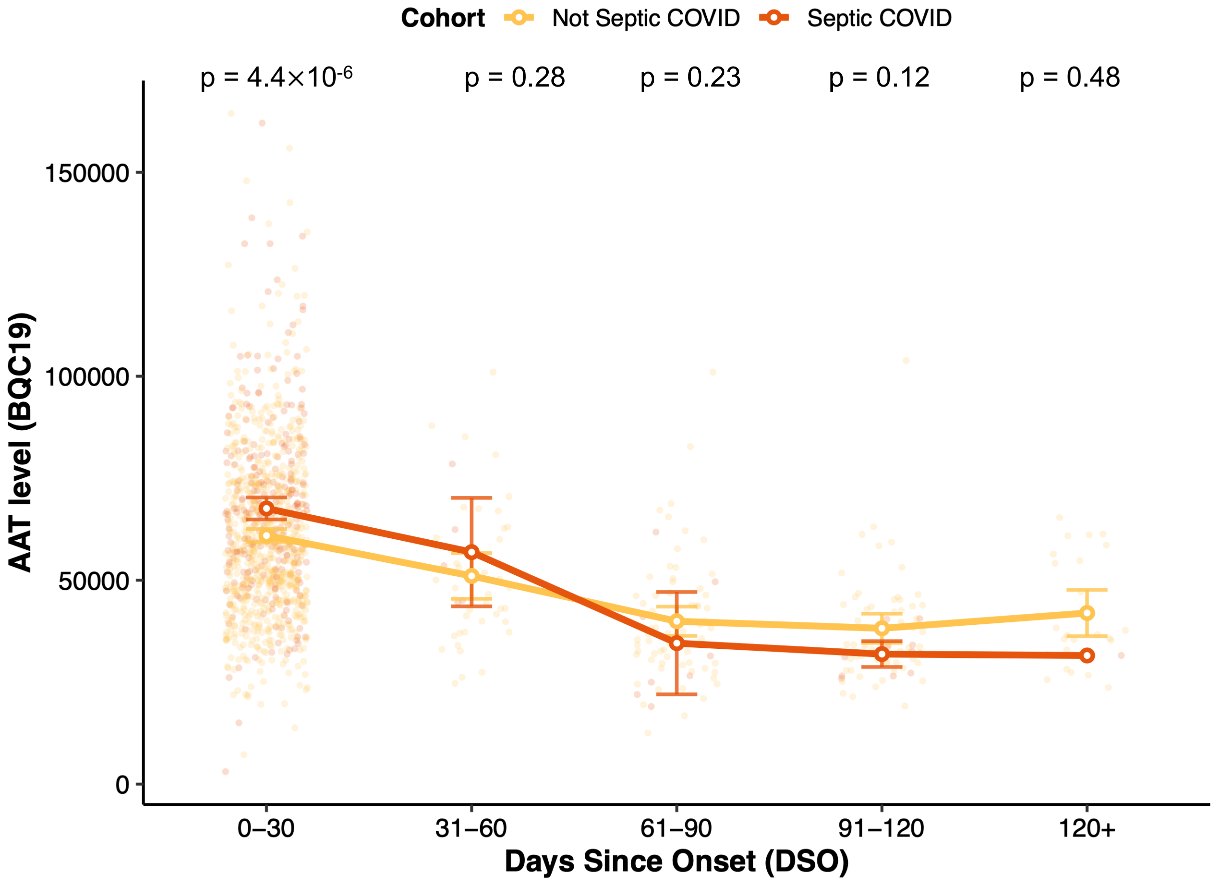


**Supplementary Figure 4. Longitudinal trajectory of AAT levels in the BQC19 cohort from symptom onset to resolution.**

Temporal evolution of circulating AAT levels quantified in the BQC19 cohort. The x-axis represents categorical time bins based on days since symptom onset (DSO): 0-30, 31-60, 61-90, 91-120, and >120 days. The y-axis represents the protein abundances. Red lines and points illustrate the trajectory of patients with septic COVID-19, while yellow lines and points denote patients with non-septic COVID. Points represent the mean protein abundance within each bin, and error bars represent the standard deviation. This longitudinal profile confirms a sharp acute-phase elevation of AAT during the first 30 days of sepsis, followed by a gradual return to physiological baseline upon clinical recovery. P-values denote significance between groups derived from the Wilcoxon rank-sum test.

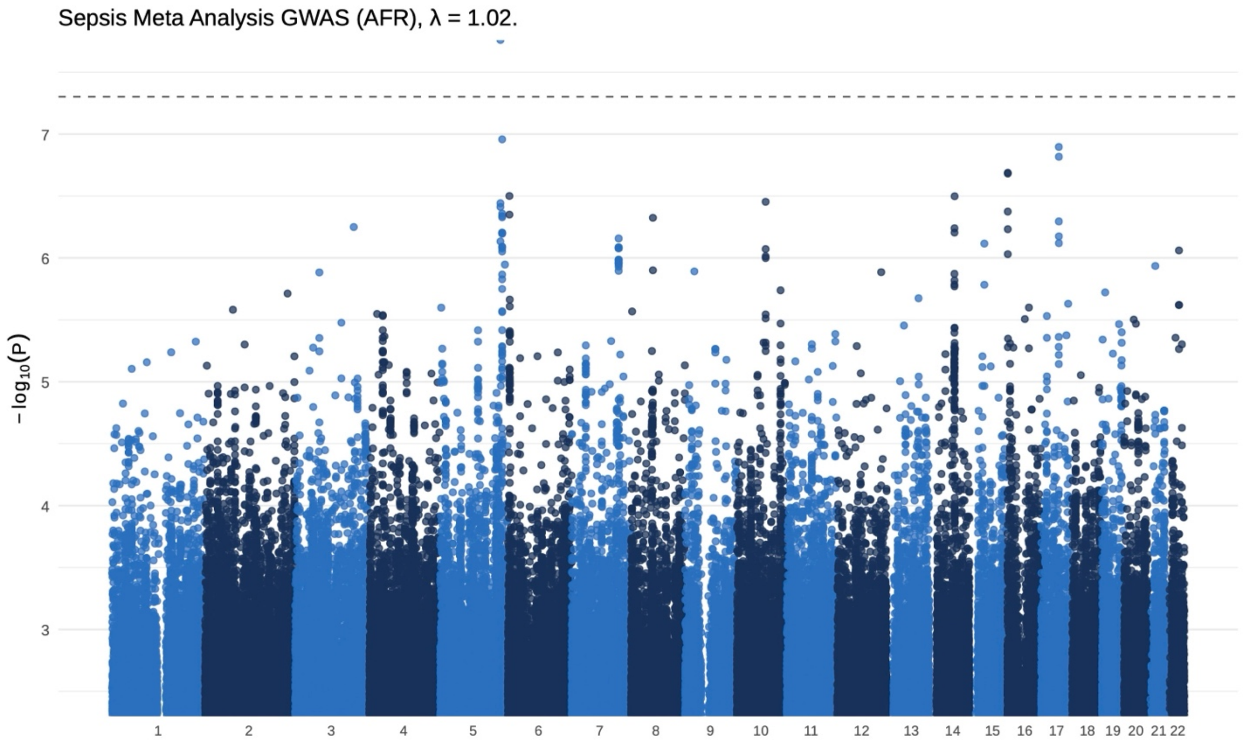


**Supplementary Figure 5. The Manhattan plot of African meta-analysis of sepsis.** Manhattan plot illustrating the meta-analysis of genetic associations with African ancestry sepsis (7,366 cases and 167,270 controls). The x-axis represents chromosomal position, and the y-axis represents the negative of the $\log_{10}(P)$ value (the higher the dot, the lower the p-value). The horizontal dashed line indicates the threshold for genome-wide significance ($P = 5\times10^{-8}$). The genomic inflation factor (λ) was 1.02.

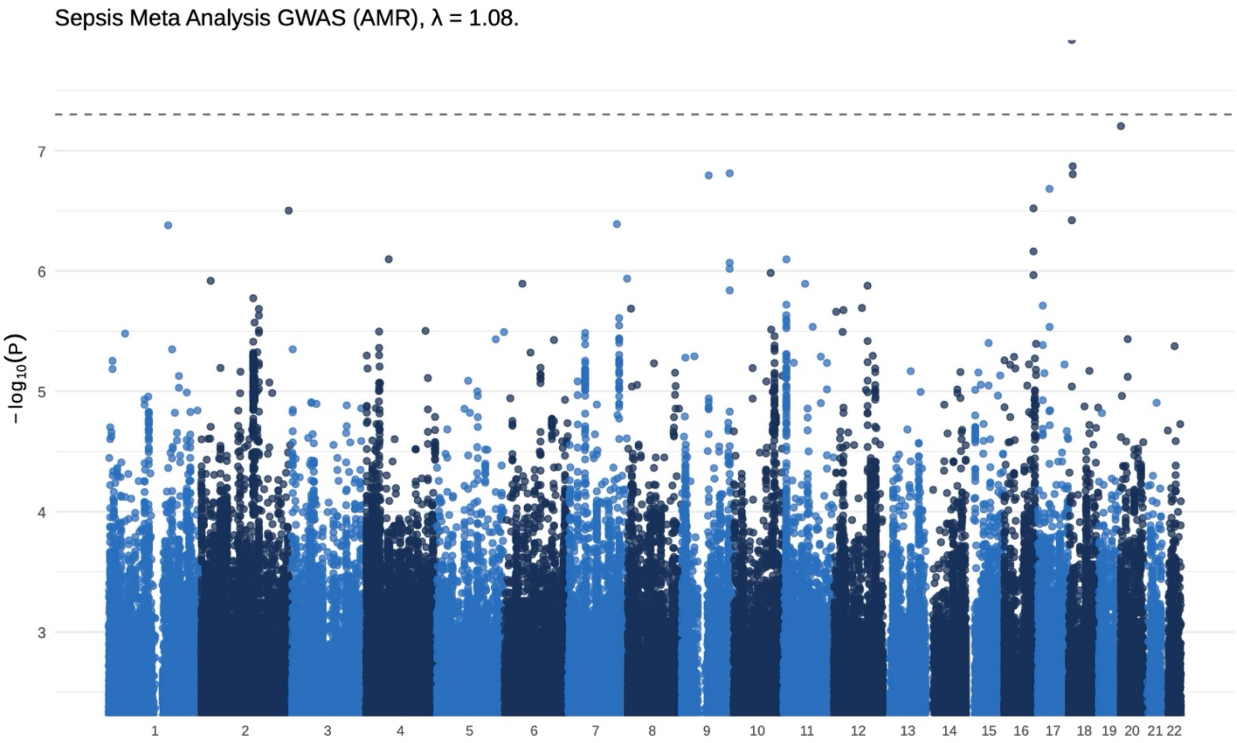


**Supplementary Figure 6. The Manhattan plot of Admixed American meta-analysis of sepsis.**

Manhattan plot illustrating the meta-analysis of genetic associations with Admixed American ancestry sepsis (6,088 cases and 103,762 controls). The x-axis represents chromosomal position, and the y-axis represents the negative of the $\log_{10}(P)$ value (the higher the dot, the lower the p-value). The horizontal dashed line indicates the threshold for genome-wide significance ($P = 5\times10^{-8}$). The genomic inflation factor (λ) was 1.08.

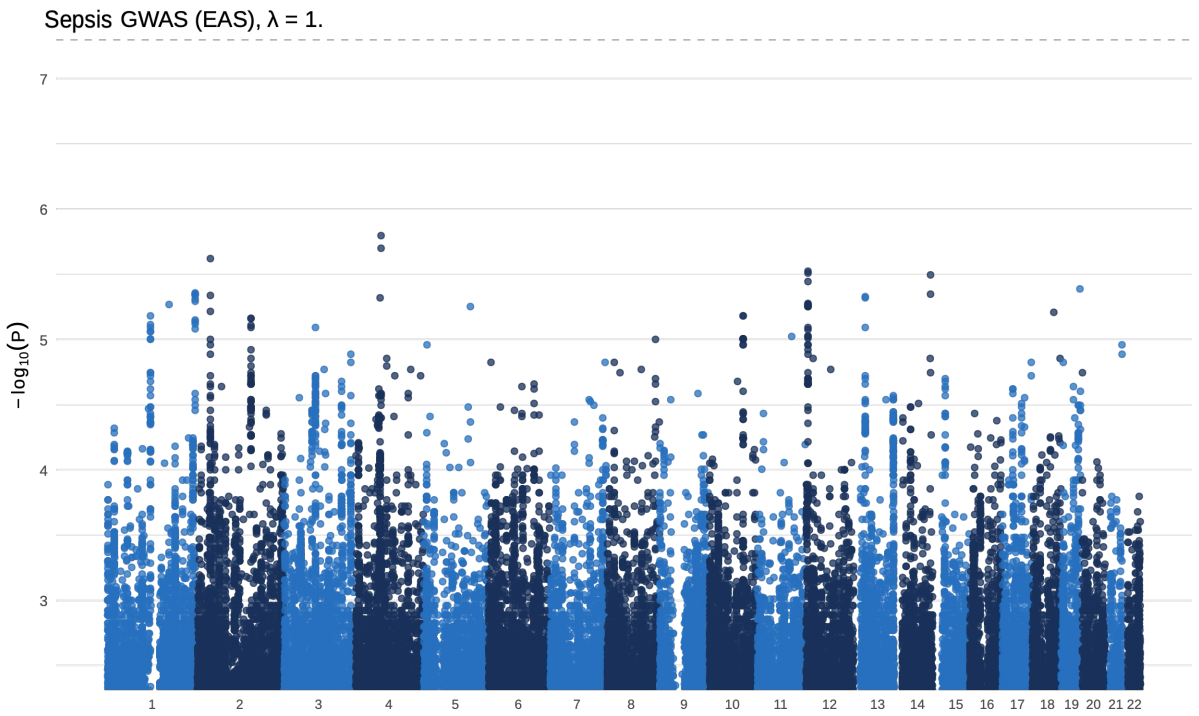


**Supplementary Figure 7. The Manhattan plot of East Asian GWAS of sepsis.** Manhattan plot illustrating the of East Asian sepsis GWAS from TPMI (2,022 cases and 339,066 controls). The x-axis represents chromosomal position, and the y-axis represents the negative of the $\log_{10}(P)$ value (the higher the dot, the lower the p-value). The horizontal dashed line indicates the threshold for genome-wide significance ($P = 5\times10^{-8}$). The genomic inflation factor (λ) was 1.

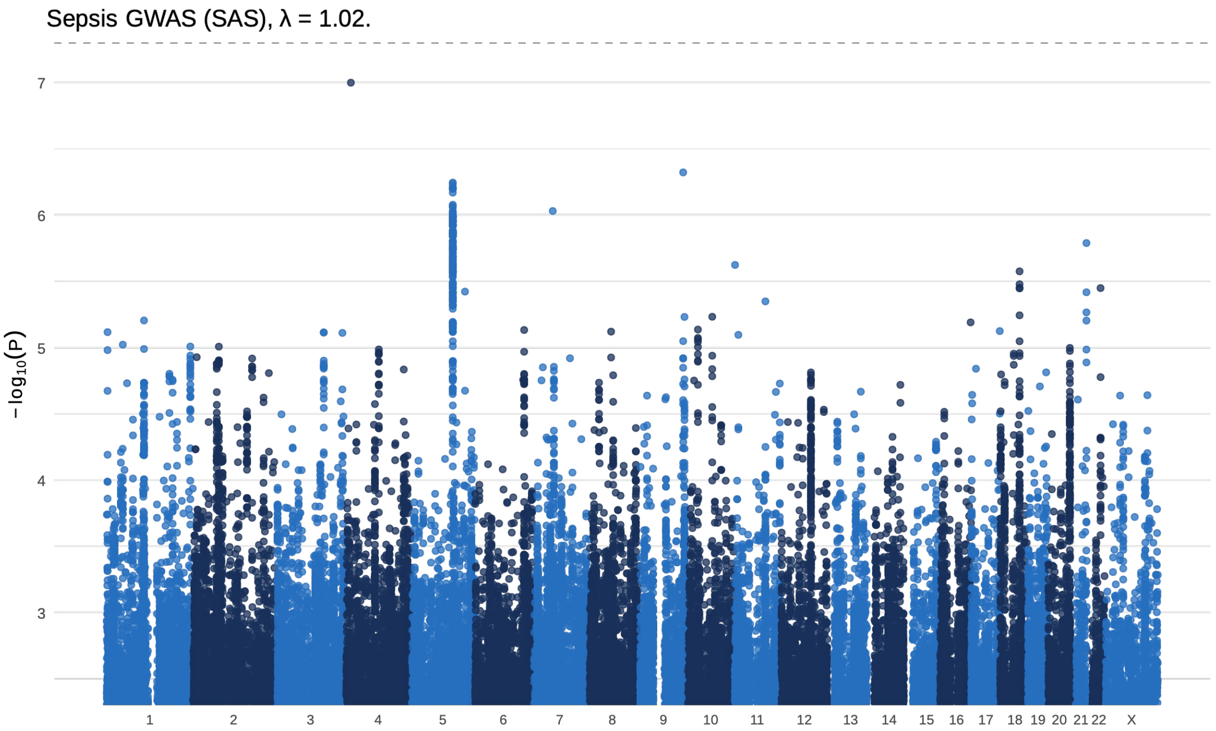


**Supplementary Figure 8. The Manhattan plot of South Asian GWAS of sepsis.** Manhattan plot illustrating the of South Asian ancestry sepsis GWAS from Genes&Health (1,146 cases and 47,343 controls). The x-axis represents chromosomal position, and the y-axis represents the negative of the $\log_{10}(P)$ value (the higher the dot, the lower the p-value). The horizontal dashed line indicates the threshold for genome-wide significance ($P = 5\times10^{-8}$). The genomic inflation factor ($\lambda$) was 1.02.
